# Spousal bereavement, mortality and risk of negative health outcomes among older adults: a population-based study

**DOI:** 10.1101/2020.04.16.20067645

**Authors:** Lucas Morin, Jonas W. Wastesson, Stefan Fors, Neda Agahi, Kristina Johnell

## Abstract

**Objective:** to examine whether spousal bereavement increases the risk of death and negative health outcomes and among older people.

**Design:** cohort study and self-controlled cohort crossover study

**Setting:** routinely collected administrative and healthcare data with individual-level linkage between several national registries in Sweden.

**Participants:** older persons (≥65 years) living in the community whose spouse died in 2013–2014, individually matched with controls.

**Main outcome measures:** death from any cause (primary outcome), acute cardiovascular events, pneumonia, hip fracture, and intentional self-harm (secondary outcomes). In the cohort study, incidence rate ratios were estimated with conditional fixed-effect Poisson regression models adjusted for potential confounders. In the self-controlled cohort crossover study, relative incidence ratios were estimated over the 12 months before and after spousal loss with unadjusted conditional fixed-effect Poisson regression including a bereavement-by-time interaction.

**Results:** 42 918 bereaved older spouses were included and matched to an equal number of married controls (mean age 78.9 [SD 7.2] years, 68% women). During the first year of follow-up, the risk of death from any cause was 1.66 (95% confidence interval 1.53 to 1.80) times higher for bereaved cases than for married controls, and bereaved cases survived on average 4.2 days shorter than married controls. Bereaved cases also experienced an increased risk of acute cardiovascular events (incidence rate ratio 1.34, 1.24 to 1.44), hip fracture (1.48, 1.30 to 1.68), pneumonia (1.14, 1.04 to 1.25), and self-harm (3.49, 2.11 to 5.76). These associations were strongly time-dependent, increasing sharply immediately after spousal loss and weakening as time elapsed. In the self-controlled cohort crossover study, the relative incidence ratios increased for all four secondary outcomes, starting already during the 6-month period preceding spousal loss.

**Conclusion:** Among older persons, the association between spousal bereavement and the risk of negative health outcomes and mortality is most likely causal. Our finding that the risk of adverse health consequences increases already during the 6 months prior to spousal loss indicates that palliative care services have an important role to play in providing timely bereavement care to spouses and other family caregivers.

**What is already known**

- Bereavement is associated with an excess risk of mortality in the surviving spouses.
- There is some evidence that spousal loss is also associated with acute cardiovascular events.
- Previous studies have often focused on young and relatively healthy groups of people, although bereavement of a spouse typically occurs as we reach older ages.

**What this study adds**

- Spousal bereavement comes with a rapid and substantial increase in mortality, acute cardiovascular events, hip fracture, pneumonia, self-harm, non-elective hospitalisation and nursing home admission.
- The excess risk of negative health events observed already during the months before spousal loss indicates the influence of an ‘anticipatory effect’.
- Although sudden spousal deaths expose surviving partners to especially high excess mortality, longer and more predictable illness trajectories do not shield spouses from the adverse health consequences of bereavement.

**Implications for clinical practice and health policy**

- Bereavement support should be offered without delay to mitigate short-term hazards and then maintained over a sufficiently long period of time.
- Such support should be provided not only to recently bereaved individuals, but also to the spouses of seriously ill older people with poor 6-month prognosis.
- Palliative care services could have an important role in providing bereavement support to older adults.

## Introduction

The loss of a spouse is a distressing life event that typically occurs in old age. Bereaved partners can be affected both psychologically and physiologically.^1,2^ This is particularly true for older adults, as spousal bereavement often marks the end of a period of high strain due to caring for a seriously ill partner. It can also represent the loss of a crucial caregiver without whom self-care can prove challenging. Evidence from census data and cohort studies suggests that bereavement is associated with a substantial excess risk of mortality and adverse health events among the surviving spouses.^3,4^ With over 1.5 million older persons experiencing spousal loss every year in the European Union, including 185 000 in the United Kingdom (**Supplementary Table 1**), the health outcomes of bereavement are an issue of high public health relevance. Despite long-standing interest ^5–13^, this phenomenon sometimes referred to as the ‘widowhood effect’ remains unclear in several aspects.

**Table 1.** Characteristics of study population at cohort entry

|  | **Bereaved cases** | **Married controls** |
| --- | --- | --- |
| **Total, No.** | 42 918 | 42 918 |
| **Sex** |  |  |
| Male | 13 581 (31.6%) | 13 581 (31.6%) |
| Female | 29 337 (68.4%) | 29 337 (68.4%) |
| **Age, years** |  |  |
| Mean, SD | 78.9 (7.2) | 78.9 (7.2) |
| 65 to 74 years | 13 944 (32.5%) | 13 960 (32.5%) |
| 75 to 84 years | 19 445 (45.3%) | 19 454 (45.3%) |
| 85 to 94 years | 9320 (21.7%) | 9286 (21.6%) |
| ≥95 years | 209 (0.5%) | 218 (0.5%) |
| **Chronic diseases** |  |  |
| Mean number, SD | 2.8 (2.7) | 2.7 (2.7) |
| Atrial fibrillation | 4675 (10.9%) | 4627 (10.8%) |
| Cerebrovascular disease | 3184 (7.4%) | 3170 (7.4%) |
| Congestive heart failure | 2779 (6.5%) | 2628 (6.1%) |
| Chronic ischemic heart disease | 7810 (18.2%) | 7632 (17.8%) |
| Chronic obstructive pulmonary disease | 2449 (5.7%) | 2230 (5.2%) |
| Dementia | 1343 (3.1%) | 1556 (3.6%) |
| Diabetes | 5891 (13.7%) | 5405 (12.6%) |
| Solid cancer | 6400 (14.9%) | 6670 (15.5%) |
| **Other conditions** |  |  |
| History of smoking-related cancer | 1540 (3.6%) | 1319 (3.1%) |
| History of alcohol-related disease | 303 (0.7%) | 159 (0.4%) |
| History of depression and mood disorders | 871 (2.0%) | 721 (1.7%) |
| History of other psychiatric diseases | 698 (1.6%) | 554 (1.3%) |
| **Hospital Frailty Risk Score** |  |  |
| Mean, SD | 2.3 (3.7) | 2.3 (3.8) |
| Low risk (<5) | 34 254 (79.8%) | 34 455 (80.3%) |
| Intermediate risk (5-9) | 6085 (14.2%) | 5874 (13.7%) |
| High risk (≥10) | 2579 (6.0%) | 2589 (6.0%) |
| **Prescription drugs** |  |  |
| Mean number of drugs, SD | 5.7 (4.1) | 5.6 (4.2) |
| Polypharmacy (≥5 drugs) | 24 285 (56.6%) | 23 686 (55.2%) |
| Any antihypertensive | 27 515 (64.1%) | 27 032 (63.0%) |
| Statins | 12 701 (29.6%) | 12 556 (29.3%) |
| VKA and platelet antiaggregants | 17 044 (39.7%) | 16 845 (39.2%) |
| **Level of education** |  |  |
| Primary education | 16 267 (37.9%) | 14 389 (33.5%) |
| Secondary education | 19 220 (44.8%) | 19 045 (44.4%) |
| Tertiary education | 6990 (16.3%) | 8572 (20.0%) |
| Missing | 441 (1.0%) | 912 (2.1%) |
| **Disposable income, thousand US dollars** |  |  |
| Median (1<sup>st</sup> and 3<sup>rd</sup> quartiles) | 27.7 (23.4 – 34.8) | 29.1 (24.1 – 38.4) |
| **Small-area level of social deprivation** |  |  |
| 1 (least deprived areas) | 9578 (22.3%) | 11 262 (26.3%) |
| 2 | 9243 (21.6%) | 9338 (21.8%) |
| 3 | 11 171 (26.0%) | 10 668 (24.9%) |
| 4 (most deprived areas) | 12 875 (30.0%) | 11 565 (27.0%) |
| Missing | 51 (0.1%) | 85 (0.1%) |

There is much debate about the nature of the relationship between spousal loss and subsequent adverse health outcomes. First, is the observed increase in the risk of death truly the *consequence* of bereavement, or is it—at least partly—attributable to phenotypical similarities between deceased individuals and their surviving spouse? For instance, assortative mating, convergence in lifestyle and shared environment throughout adulthood may very well be at play.^14^ Previous studies have often been unable to convincingly establish a causal link between bereavement and mortality because of a lack of information on socioeconomic and health-related confounders at the time of spousal loss.^3^ Second, is the potential effect of bereavement mainly fuelled by acute factors that influence health immediately after spousal loss or is it a mostly silent and prolonged process that accumulates over time? Although there is some evidence that the mortality risk associated with the loss of a partner is highest during the first year after widowhood and becomes weaker as time elapses, little is known about the exact shape of this association over time.^3,15,16^ Third, does the potential effect of spousal bereavement on physical health extend beyond mortality? Prior research has shown that compared with non-bereaved individuals those who recently lost a spouse may have a higher risk of acute cardiovascular events.^17^ However, the only large study published to date on this topic ^18^ relied exclusively on electronic records from primary care without linkage to hospital diagnoses or causes of death, thereby potentially underestimating the actual number of cases. Fourth, most studies have examined the association between widowhood and mortality without differentiating the end-of-life trajectory of the deceased spouse. For instance, are older persons whose spouse died from dementia (often preceded by a long period of informal care) at greater excess risk of negative health consequences than those who lost their partner from another cause of death? Investigating potential differences between illness trajectories is an important step to identify caregivers at high risk of adverse health events and to design targeted interventions. Finally, there is a need to evaluate health outcomes longitudinally, both *before* and *after* spousal loss. Measuring within-individual changes in the risk of experiencing negative health outcomes would allow for disentangling the potential effect of bereavement from that of pre-bereavement experiences such as caring for a seriously ill spouse near the end of life.^19–21^

The present study aimed to overcome some of the limitations of previous work by addressing each of these five issues. We examined the risk of death and adverse health outcomes associated with spousal bereavement among community-dwellers aged 65 years and older by analysing routinely collected administrative and healthcare data with two different epidemiological designs (matched cohort and self-controlled cohort crossover) and by applying a variety of methods to assess and mitigate the risk of bias inherent to the observational nature of the study.

## Methods

### Data source

We used routinely collected data with individual-level linkage between several national registers in Sweden.^22,23^ The database used for this study contains pseudonymised information about the sociodemographic characteristics, marital status, medical diagnoses, drug prescriptions, hospitalisations, nursing home admissions, and vital status of all older adults aged 65 years and over (see **Supplementary Table S2** for detailed information). Married individuals are linked to their respective spouse through a unique identifier, which is updated continuously to reflect changes in marital status.

### Study population

The study population included people aged 65 years or over registered in Sweden between 1 January 2013 and 31 December 2014. A total of 50 058 married older adults who died from any cause during that period were identified in the National Cause of Death Register ^24^, of which 47 682 (95.3%) could be linked to their surviving spouse. Among these bereaved individuals, we selected those who met the following eligibility criteria: (1) they were aged 65 years or over at the time of spousal loss; (2) they were community-dwellers (i.e. not living in nursing homes or residential care facilities); and (3) they had not already experienced spousal bereavement during the previous 5 years. In addition, spouses who died on the exact same day or from shared causes within 30 days (e.g. as a consequence of the same road-traffic accident) were excluded. To avoid misclassification of the timing of the exposure, we compared the date of spousal loss to the date of change in civil status for the surviving spouse reported to the Swedish Tax Agency, and we found a 99.96% agreement. A total of 42 918 bereaved older adults were thereafter included in the analyses (**Figure 1**).

**Figure 1.**
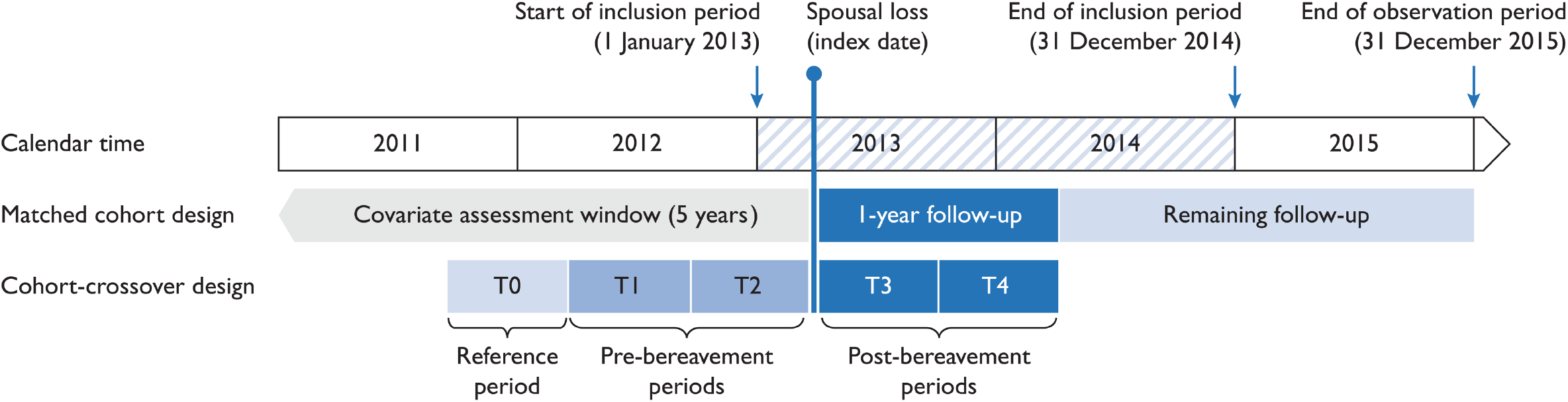
Illustration of matched cohort and cohort-crossover designs. In the matched cohort design, each bereaved older adult (case) is matched on sex and age to a single married control, without replacement. Follow-up starts at the index date, i.e. the date of spousal loss for the cases, and the matched date for the controls. Individuals are followed until the event of interest, death, or the end of the observation period. In the cohort-crossover design, individuals serve as their own controls over time. Within-person change in risk of adverse event is calculated by comparing the event rates at baseline (T0, 547–366 days before index date) to event rates during pre-bereavement (T1, 365–184 days and T2, 183–1 days before index date) and post-bereavement periods (T3, 1–183 days and T4: 184–365 days after index date). For each period from T1 to T4, within-person change among bereaved cases is then compared to the within-person change among married controls.

Bereaved individuals were matched 1:1 by sex and age (1-month bands) to a control group of married older adults sampled from the Total Population Register using the date of spousal loss among the bereaved as index date. Married individuals in the risk set were eligible as controls if they did not experience spousal loss during the 5 years before or during the year after the index date, and if they were not already living in nursing homes at the time of cohort entry. Matching was done without replacement, so that each married control was assigned to a single bereaved case.

### Outcomes

The primary outcome was death from any cause after spousal loss, which was assessed from the National Cause of Death Register. Secondary outcomes included acute cardiovascular events (composite of acute myocardial infarction, other acute coronary syndrome, pulmonary embolism, acute pericarditis, acute myocarditis, cardiac arrest, stroke, and aortic aneurysm or dissection), pneumonia, hip fracture, and intentional self-harm. These outcomes were defined by the first occurrence of either an emergency department visit, a non-elective hospitalisation or death due to specific causes identified with International Statistical Classification of Diseases 10^th^ Revision (ICD-10) codes. The complete list of codes is available in **Supplementary Table S3**. We also considered two secondary outcomes related to healthcare utilisation, namely non-elective hospitalisations for any cause and nursing home admission.

### Study design

We used both matched cohort and self-controlled crossover designs.

#### Cohort study

The risk of mortality and adverse health outcomes during the year after spousal loss was compared between bereaved older adults and their matched married counterparts. For the main analysis, we considered a 1-year period to be the most clinically relevant based on previous reports showing that the effect of bereavement was often rapid, ^1,3,15,18^ but we decided *a priori* to also report health outcomes during the entire available observation time (namely, up to 31 December 2015 with a median follow-up time of 1.9 years). The burden of chronic diseases ^25^ and the Hospital Frailty Risk Score ^26^ at baseline were assessed by applying validated algorithms based on data reported during the 5 years before spousal loss (**Supplementary Table S4**). Prior history of smoking-related cancer, alcohol-related diseases and mental health problems was captured in the National Patient Register for the same period of time (**Supplementary Table S5**). We analysed medication use at baseline by using information from the Swedish Prescribed Drugs Register.^27^ In addition, the socioeconomic position of study participants was described by using routinely collected administrative data about their highest educational attainment, their equivalised disposable income during the year before spousal loss, as well as a composite measure of the area level of social deprivation (**Supplementary Table S6**).

#### Self-controlled cohort crossover study

In self-controlled studies, cases are used as their own controls to eliminate the risk of bias due to between-individual unmeasured confounders.^28–31^ King and colleagues recently conducted a self-controlled case series analysis to examine the mortality risk after the loss of a partner compared with before.^32^ However, since it can be argued that the experience of bereavement is not a transient exposure clearly defined in time, these methods can yield biased estimates.^33^ Instead, we used a cohort crossover design combining the self-controlled and the cohort approaches to measure the within-individual change in the risk of experiencing adverse health outcomes before and after spousal loss. This approach has, for instance, been proposed to study the risk of aortic dissection during pregnancy ^34^ or to quantify the excess risk of injury during the period shortly before and after a diagnosis of cancer.^35^ In our study, we defined a reference period from 18 until 12 months before spousal loss (T0), and we calculated the relative risk of adverse health outcomes from 1 year before until 1 year after spousal loss by dividing person time into four 6-month periods (182 days), from T1 to T4 (**Figure 2**). By design, the relative risk of fatal events during the pre-bereavement period cannot be calculated.

**Figure 2.**
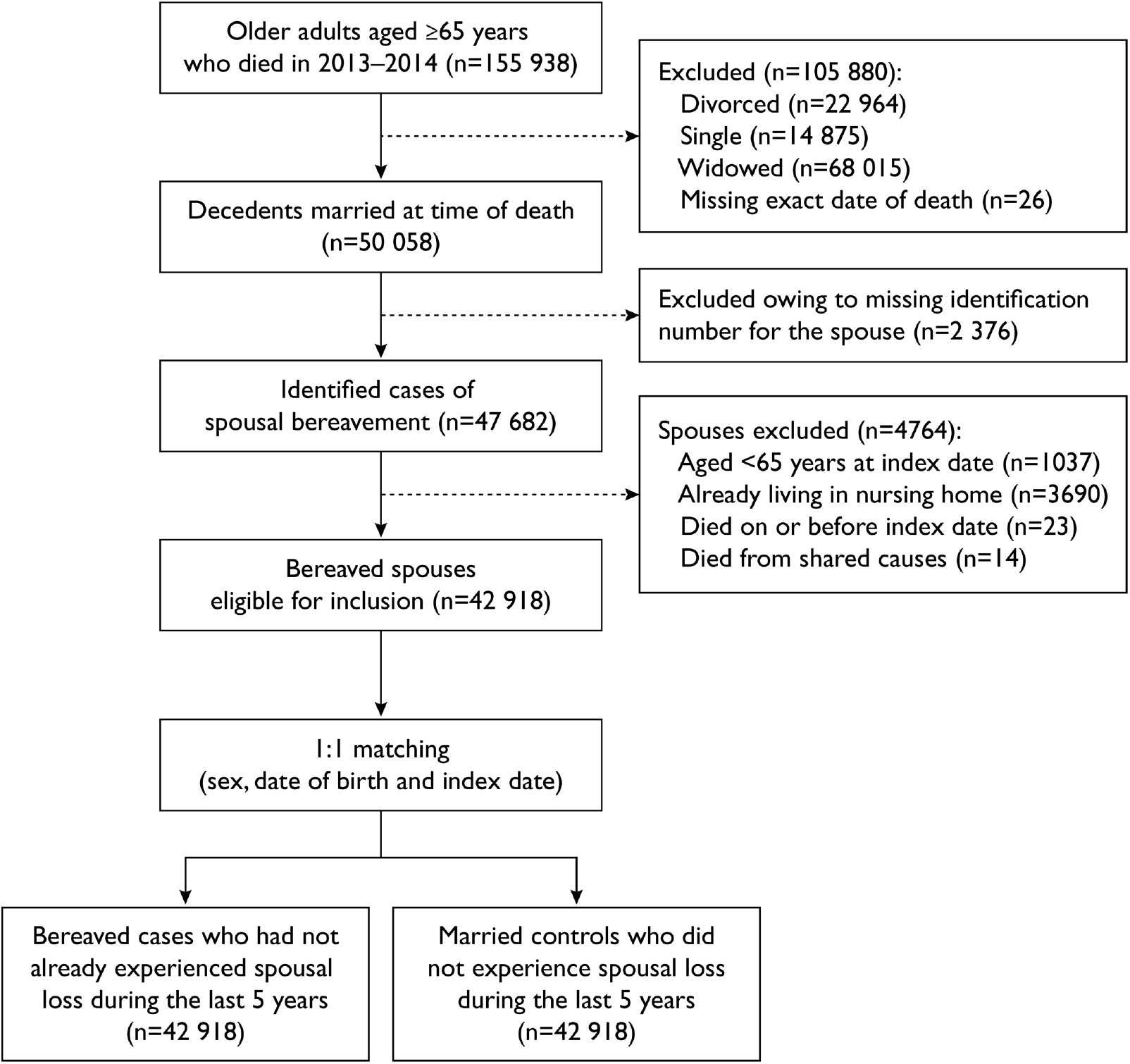
Flow diagram of matched cohorts.

### Statistical analysis

#### Matched cohort study

Summary statistics were calculated to describe the characteristics of cases and controls at baseline. We used conditional fixed-effect Poisson regression models to compare the incidence rate of death and adverse health outcomes between cases and controls.^36–38^ Incidence risk ratios (IRR) were conditioned on the matched set (sex and age) and were further adjusted on relevant confounders, which were selected for each outcome separately considering the imbalance of covariates at baseline (>0.5% difference) and subject matter knowledge. Individuals were followed from the index date until the earliest of the first event date, a censoring event (death or emigration), or the end of observation time. As previous reports suggested that spousal loss may have more detrimental effects on health among men than among women ^3^, all analyses were stratified by sex. To overcome some of the caveats of relative summary metrics of between-group survival differences,^39^ we compared the adjusted restricted mean survival time of bereaved and married individuals.^40^ In addition, we assessed the time-dependent effect of spousal loss on mortality by computing piecewise Poisson regression models and by fitting flexible parametric survival models with restricted cubic splines to plot a smooth curve of the hazard ratio as a function of time since bereavement.^41^ We also investigated variations in the hazard ratio for mortality throughout the age span and across percentiles of income by constructing restricted cubic splines for these two continuous variables.

#### Self-controlled cohort crossover study

We used conditional fixed-effect Poisson regression models to compare the incidence rate of non-fatal events during the pre-bereavement and post-bereavement periods with the incidence rate during the reference period (18 to 12 months before spousal loss). For each hazard period, person-time was calculated from the beginning of the interval until either the first event of interest, the date of death or emigration, or the end of the interval. We computed relative incidence ratios (RIR) for each outcome by entering a bereavement-by-time interaction term in the model.^42^ This was done to account for the fact that incidence rates during pre-bereavement periods are conditional on survival until the date of spousal loss and, thus, that the observed incidence rates after that date may partly stem from left-truncation bias. Relative incidence ratios can be interpreted as the relative increase in the risk of experiencing adverse health outcomes among bereaved cases compared with the increase among married controls. This analytical strategy presents the advantage of eliminating the influence of time-invariant confounders while also removing the effect of time trends in the events.^43^ We obtained 95% confidence intervals (CIs) by using robust standard errors to account for clustering within individuals. Analyses were performed with SAS JMP version 13 (SAS Institute Inc., Cary, NC) and Stata version 14 (StataCorp., College Station, TX).

### Sensitivity analyses

We conducted five types of prespecified sensitivity analyses. First, we compared the incidence rate ratios obtained from adjusted conditional fixed-effect Poisson regression models (main analysis) with the results from two alternative analyses: unconditional Poisson regression models (further adjusted for sex and age) and Cox proportional hazards regression models stratified on the matched pairs. Second, we assembled a set of controls who were neither married nor recently bereaved. Because single and divorced individuals are too few in these birth cohorts and tend to be highly selected in terms of socioeconomic position and lifestyle, we used a cohort of long-time widows and widowers. Each bereaved case was matched on sex and age to a widowed person who had experienced spousal loss more than 5 years prior to the index date (median 11 years) and who did not remarry afterwards. We hypothesised that if the causal effect of spousal bereavement is mostly acute and weakens as time elapses, then the excess risk of adverse health outcomes should be substantially greater among newly bereaved cases compared to married individuals than compared to long-time widows and widowers. Third, we made use of *negative control outcomes*, namely conditions thought to be unrelated to bereavement and thus presumed to have null effect sizes,^44,45^ to unravel potential sources of bias due to unmeasured confounders related to shared lifestyle and environmental factors. These negative control outcomes include the incidence of solid cancer, hallux valgus, Parkinson’s disease and cataract during the first year of follow-up among individuals without these conditions at baseline. We reasoned that if the association between spousal loss and subsequent health events was truly causal, no association should be observed with biologically implausible health outcomes such as the risk of developing a new solid tumour or Parkinson’s disease within one year.^46^ A positive association would thus indicate the existence of residual confounding. Fourth, we calculated the E-value for each of the outcomes observed in our matched cohort design. This methodology has been proposed by VanderWeele and colleagues as a way to evaluate how susceptible an association is to potential unmeasured or uncontrolled confounding.^47^ The E-value can be defined as the minimum strength of association that an unmeasured confounder would need to have with both the exposure and the outcome to fully explain away the observed association between them, conditional on the measured covariates. Finally, we expanded the definition of acute cardiovascular events to include a broader range of conditions than the ones included in the main analysis (e.g. acute endocarditis, non-rheumatic valve disorders, cardiomyopathies, deep vein thrombosis) and we extended the definition of fall-related injuries beyond hip fracture.

### Post-hoc analyses

We also performed three sets of post-hoc analyses, which were not prespecified. First, we considered the possibility that an increased risk of adverse health outcomes among the bereaved may be partially due to the experience of caring for a seriously ill spouse nearing the end of life. To investigate this potential mechanism, we stratified the main analysis in four distinct subgroups according to the illness trajectory of the deceased spouse: (i) cancer, (ii) organ failure, (iii) dementia and neurodegenerative disorders, and (iv) sudden death. Individuals were assigned to a single illness trajectory by using a methodology described elsewhere.^48^ Second, we compared the effect estimate of spousal bereavement on subsequent mortality according to the Hospital Frailty Risk Score at baseline to assess whether the excess risk of death was observed only among the frailest and most vulnerable individuals or not. Finally, we considered the competing risk of death in the incidence of adverse health outcomes.

### Ethical approval

The Regional Ethical Review Board in Stockholm approved the study. The need for individual consent was waived as the study relies exclusively on pseudonymised administrative and healthcare data.

## Results

### Study population

We identified a total of 42 918 older adults who experienced spousal loss during the study period and met our inclusion criteria, matched 1:1 to an equal number of married controls. The proportion of women in each group was 68.4% and the mean age was 78.9 (SD 7.2) years. We identified 18 and 9 persons in same-sex relationships among bereaved cases and married controls, respectively. Baseline characteristics were similar across groups (**Table 1**), although bereaved older adults were more likely to have a history of diabetes (13.7% vs. 12.6%), to be prescribed ≥5 concurrent drugs (56.6% vs. 55.2%), to have a lower level of education (37.9% vs. 33.5%), and to be living in more socially deprived areas (30.0% vs. 27.0%) than their married counterparts. They were also more likely to have a prior history of smoking-related cancer (3.6% vs. 3.1%), alcohol-related disease (0.7% vs. 0.4%) or depression and mood disorders (2.0% vs 1.7%). During follow-up, 87 bereaved cases and 48 married controls emigrated from Sweden. The characteristics of long-time widowed controls (n=42 918) were found to be similar to that of the bereaved cases, as reported in **Supplementary table S7**.

#### Matched cohort study

Table 2 summarises the occurrence of deaths and adverse health events after spousal loss. During the first year of follow-up, 2060 bereaved cases and 1306 married controls died (adjusted IRR 1.66, 95% CI 1.53–1.80). Unadjusted and adjusted survival curves are presented in **Supplementary Figure S1**. Adjusted restricted mean survival time analyses showed that bereaved cases survived on average 4.2 days shorter than married controls when they were followed for 1 year, and 20.5 days shorter when follow-up was extended to 3 years (**Supplementary table S8**). We also observed an increased risk of acute cardiovascular events (IRR 1.34, 95% CI 1.24–1.44), hip fracture (IRR 1.48, 95% CI 1.30–1.68), pneumonia (1.14, 95% CI 1.04–1.25), and self-harm (IRR 3.49, 95% CI 2.11–5.76) among bereaved cases compared with their married counterparts. Although effect estimates decreased for all outcomes when we considered the entire available follow-up time, the association with spousal loss remained (IRR for all-cause mortality 1.36, 95% CI 1.29–1.43). The excess risk of mortality was highest during the first 3 months after spousal loss, and gradually decreased throughout the remaining follow-up (**Supplementary Figure S2 and Table S9**). We observed a similar pattern for the time-varying association of spousal loss with adverse health events (**Supplementary Figure S3**). As shown in **Table 3**, bereaved older adults also had higher incidence rates of non-elective hospitalisations (IRR 1.17, 95% CI 1.14–1.21) and nursing home admissions (IRR 3.10, 95% CI 2.81–3.45).

**Table 2.** Incidence rate ratios and 95% confidence intervals for all-cause mortality and adverse health events after spousal loss (matched cohort analysis)

|  | First year of follow-up |  | Complete follow-up |  |
| --- | --- | --- | --- | --- |
|  | Bereaved cases | Married controls | Bereaved cases | Married controls |
| **All-cause mortality** |  |  |  |  |
| No. of deaths | 2060 | 1306 | 4191 | 3229 |
| Person years of follow-up | 41 866 | 42 359 | 81 671 | 83 285 |
| Rate per 1000 person-years (95% CI) | 49.2 (47.1–51.4) | 30.8 (29.2–32.5) | 51.3 (49.8–52.9) | 38.8 (37.5–40.1) |
| Unadjusted incidence rate ratio (95% CI)<sup>a</sup> | 1.63 (1.52–1.75) |  | 1.37 (1.30–1.43) |  |
| Adjusted incidence rate ratio (95% CI)<sup>b</sup> | 1.66 (1.53–1.80) |  | 1.36 (1.29–1.43) |  |
| **Acute cardiovascular event** |  |  |  |  |
| No. of persons with the event | 1825 | 1395 | 3396 | 2772 |
| Person years of follow-up | 41 259 | 41 848 | 79 481 | 81 378 |
| Rate per 1000 person years (95% CI) | 44.2 (42.2–46.3) | 33.3 (31.6–35.1) | 42.7 (41.3–44.2) | 34.1 (32.8–35.4) |
| Unadjusted incidence rate ratio (95% CI)<sup>a</sup> | 1.34 (1.24–1.43) |  | 1.26 (1.20–1.33) |  |
| Adjusted incidence rate ratio (95% CI)<sup>c</sup> | 1.34 (1.24–1.44) |  | 1.25 (1.18–1.32) |  |
| **Hip fracture** |  |  |  |  |
| No. of persons with the event | 645 | 449 | 1241 | 944 |
| Person years of follow-up | 41 551 | 42 125 | 80 520 | 82 446 |
| Rate per 1000 person years (95% CI) | 15.5 (14.4–16.8) | 10.7 (9.7–11.7) | 15.4 (14.6–16.3) | 11.4 (10.7–12.2) |
| Unadjusted incidence rate ratio (95% CI)<sup>a</sup> | 1.44 (1.27–1.62) |  | 1.32 (1.21–1.44) |  |
| Adjusted incidence rate ratio (95% CI)<sup>d</sup> | 1.48 (1.30–1.68) |  | 1.34 (1.23–1.47) |  |
| **Pneumonia** |  |  |  |  |
| No. of persons with the event | 1155 | 1000 | 2153 | 1999 |
| Person years of follow-up | 41 410 | 41 949 | 80 086 | 81 808 |
| Rate per 1000 person years (95% CI) | 27.9 (26.3–29.5) | 23.8 (22.4–25.4) | 26.9 (25.8–28.0) | 24.4 (23.4–25.5) |
| Unadjusted incidence rate ratio (95% CI)<sup>a</sup> | 1.17 (1.07–1.27) |  | 1.09 (1.02–1.16) |  |
| Adjusted incidence rate ratio (95% CI)<sup>e</sup> | 1.14 (1.04–1.25) |  | 1.09 (1.02–1.16) |  |
| **Self-harm** |  |  |  |  |
| No. of persons with the event | 89 | 30 | 150 | 66 |
| Person years of follow-up | 41 803 | 42 316 | 81 560 | 83 233 |
| Rate per 1000 person years (95% CI) | 2.1 (1.7–2.6) | 0.7 (0.5–1.0) | 1.8 (1.6–2.2) | 0.8 (0.6–1.0) |
| Unadjusted incidence rate ratio (95% CI)<sup>a</sup> | 2.97 (1.96–4.49) |  | 2.27 (1.70–3.04) |  |
| Adjusted incidence rate ratio (95% CI)<sup>f</sup> | 3.49 (2.11–5.76) |  | 2.41 (1.73–3.35) |  |

<sup>a</sup> *Unadjusted conditional Poisson regression model with grouping on the matched sets.*
<sup>b</sup> *Conditional Poisson regression model with grouping on the matched sets (sex and age), adjusted for chronic obstructive pulmonary disease (COPD), diabetes, chronic heart failure, cerebrovascular disease, history of ischemic heart diseases, history of solid cancer in the previous 5 years, number of other chronic comorbidities at baseline, and level of education.*
<sup>c</sup> *Conditional Poisson regression model with grouping on the matched sets (sex and age),, adjusted for diabetes, chronic heart failure, history of ischemic heart disease, history of cerebrovascular disease, atrial fibrillation, cardiac valve disease, peripheral vascular disease, other cardiovascular disease, number of other chronic comorbidities at baseline, and level of education.*
<sup>d</sup> *Conditional Poisson regression model with grouping on the matched sets (sex and age), adjusted for osteoporosis, osteoarthritis, Parkinson's disease, dementia, number of other chronic comorbidities at baseline, history of fall-related injury and history of alcohol-related disease in the past 5 years.*
<sup>e</sup> *Conditional Poisson regression model with grouping on the matched sets (sex and age), adjusted for diabetes, COPD, asthma, other chronic respiratory disease, autoimmune disease, and number of other chronic comorbidities at baseline.*
<sup>f</sup> *Conditional Poisson regression model with grouping on the matched sets (sex and age), adjusted for depression and other mood disorders, other psychiatric diseases (e.g. addictions, schizophrenia), solid cancer, and number of other chronic comorbidities at baseline.*

**Table 3.** Incidence rate ratios and 95% confidence intervals for non-elective hospitalisations and nursing home admissions after spousal loss (matched cohort analysis)

|  | First year of follow-up |  | Complete follow-up |  |
| --- | --- | --- | --- | --- |
|  | Bereaved cases | Married controls | Bereaved cases | Married controls |
| **Non-elective hospitalisation** |  |  |  |  |
| No. of persons with the event | 14 697 | 13 139 | 21 650 | 20 114 |
| Person years of follow-up | 33 989 | 35 367 | 56 823 | 60 296 |
| Rate per 1000 person-years (95% CI) | 432.4 (425.5–439.5) | 371.5 (365.2–377.9) | 381.0 (376.0–385.1) | 333.6 (329.0–338.2) |
| Unadjusted incidence rate ratio (95% CI)<sup>a</sup> | 1.17 (1.14–1.20) |  | 1.14 (1.12–1.17) |  |
| Adjusted incidence rate ratio (95% CI)<sup>b</sup> | 1.17 (1.14–1.21) |  | 1.15 (1.12–1.17) |  |
| **Nursing home admission** |  |  |  |  |
| No. of persons with the event | 2283 | 894 | 3554 | 1988 |
| Person years of follow-up | 41 515 | 42 476 | 78 218 | 81 789 |
| Rate per 1000 person-years (95% CI) | 55.0 (52.8–57.3) | 21.0 (19.7–22.5) | 45.4 (44.0–47.0) | 24.3 (23.3–25.4) |
| Unadjusted incidence rate ratio (95% CI)<sup>a</sup> | 2.70 (2.49–2.92) |  | 1.95 (1.84–2.06) |  |
| Adjusted incidence rate ratio (95% CI)<sup>c</sup> | 3.11 (2.81–3.45) |  | 2.14 (2.00–2.30) |  |

<sup>a</sup> Unadjusted conditional Poisson regression model with grouping on the matched sets.
<sup>b</sup> Conditional fixed-effect Poisson regression model with grouping on the matched sets (sex and age), adjusted for hospital frailty risk score, chronic obstructive pulmonary disease, diabetes, chronic heart failure, cerebrovascular disease, history of ischemic heart diseases, history of solid cancer in the previous 5 years, number of other chronic comorbidities at baseline, and level of education.
<sup>c</sup> Conditional fixed-effect Poisson regression model with grouping on the matched sets (sex and age), adjusted for hospital frailty risk score, dementia, Parkinson's disease, chronic obstructive pulmonary disease, diabetes, chronic heart failure, cerebrovascular disease, number of other chronic comorbidities at baseline, and level of education.

In subgroup analyses, we found that the association between bereavement and 1-year mortality was higher among men (IRR 1.78, 95% CI 1.59–2.01) than among women (IRR 1.59, 95% CI 1.42–1.79). Men also had a greater excess risk of acute cardiovascular events than women, but a smaller excess risk of hip fracture and self-harm (**Supplementary Table S10**). The excess risk of non-elective hospitalisation and nursing home admission was similar among men and women (**Supplementary Table S11**). For both men and women, the estimated hazard ratio for death during the first year after spousal loss peaked between the ages of 65 and 75 years (**Supplementary Figure S4**). We found no evidence of a scientifically important association between the level of income during the year before spousal loss and the hazard ratio for death during the first year after spousal loss (**Supplementary Figure S5**). The excess risk of mortality associated with bereavement was similar in magnitude across all levels of small-area social deprivation (**Supplementary Table S12**).

### Self-controlled cohort crossover study

**Figure 3** shows the relative incidence ratio (RIR) for acute cardiovascular events, hip fractures, pneumonia and self-harm during the different hazard periods before and after spousal loss, compared with the reference period T0 (i.e. 18 to 12 months before spousal loss). We observed an increase in the relative incidence ratio for all four outcomes, starting around 6 months before spousal loss and reaching a peak during the 6 months after. Hence, while between T0 and T3 the incidence rate of acute cardiovascular events increased from 20.3 to 36.1 per 1000 person-years among the bereaved cases, it increased from 20.6 to 27.8 per 1000 person-years among the married controls (RIR 1.34, 95% CI 1.12– 1.60). Detailed results are available in **Supplementary Table S13**. While the RIR for hip fractures remained statistically non-significant between T1 and T4 because of a limited number of events during each hazard period, extending the analysis to all fall-related hospitalisations led to similar point estimates but with substantially narrower 95% confidence intervals (**Supplementary Table S14**).

**Figure 3.**
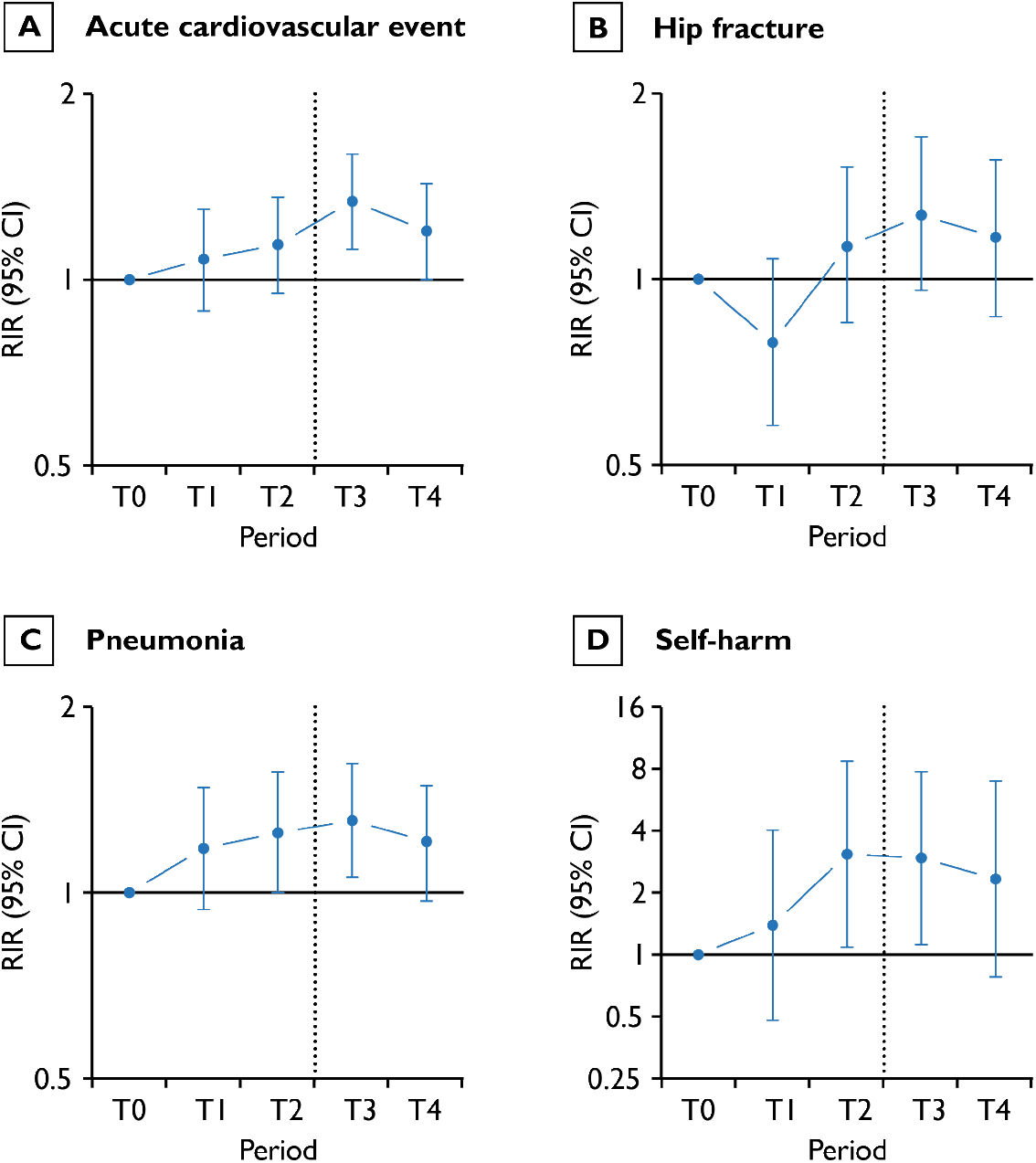
Relative incidence ratios and 95% confidence intervals for adverse health events during the year before and the year after spousal loss (self-controlled cohort crossover analysis) Abbreviations: RIR, Relative incidence ratio; CI, confidence interval Hazard periods from T0 to T4 last 6 months each, starting from 18 months before the index date and ending 12 months after the index date. Point estimates and 95% confidence intervals are provided in Supplementary Table S13.

### Sensitivity analyses

In the matched cohort study, the effect estimates calculated in our main analysis did not change in either direction or magnitude when using alternative statistical models (**Supplementary Table S15**). Compared with long-time widowed controls, newly bereaved spouses had a substantially higher risk of death, adverse health event, non-elective hospitalisation and nursing home admission, albeit with smaller effect sizes than when compared with married controls (**Supplementary Tables S16 and S17**). We found no evidence of an association between spousal loss and the risk of incident solid cancer, hallux valgus, Parkinson’s disease or cataract diagnoses, which were considered as negative control outcomes in the present study (**Supplementary Tables S18-19 and Supplementary Figure S6**). The E-values calculated for each outcome under different modelling assumptions show that our adjusted effect estimates could only be explained away by unmeasured confounding of substantial magnitude. For instance, the observed incidence rate ratio for all-cause mortality of 1.66 could be fully explained away by an unmeasured confounder that was associated with both spousal loss and risk of death by a risk ratio of 2.70 each, above and beyond the measured confounders (**Supplementary Table S20**). As shown in **Supplementary Table S21**, we found that the excess risk of acute cardiovascular events associated with spousal loss was particularly high for acute stroke (IRR 1.36, 95% CI 1.21–1.53), pulmonary embolism (IRR 1.31, 95 % CI 1.06–1.62) and aortic aneurysm or dissection (IRR 1.79, 95% CI 1.27–2.52), but only modest for acute myocardial infarction (IRR 1.14, 95% CI 1.00–1.30). Extending the definition of acute cardiovascular events to a broader range of conditions did not change our main findings (**Supplementary Table S22**). Similarly, although extending the scope of fall-related injuries beyond hip fractures led to a substantial increase in the number of events and to a slight reduction of the effect size of the observed association with bereavement (**Supplementary Table S23**), the results remained within the same order of magnitude as in the main analysis.

### Post-hoc analyses

To consider the possibility that the observed increase in poor health outcomes among bereaved older adults may be partially due to the experience of caring for a seriously ill spouse nearing the end of life, we stratified the main analysis according to the presumed illness trajectory of the deceased spouse. Older adults whose spouse died from a sudden cause of death had the greatest excess risk of death and hip fracture compared with their married counterparts, while those who lost a spouse from cancer had the greatest excess risk of pneumonia. Moreover, individuals whose spouse died from organ failure had a higher-than-average excess risk of self-inflicted injuries and suicide (**Supplementary Table S24**). In subgroup analyses, we found that, while the risk of death was substantially higher among individuals with a high Hospital Frailty Risk Score at baseline, the relative difference between bereaved cases and married controls was largest among those at intermediate risk of frailty (**Supplementary Table S25 and Supplementary Figure S7**). There was no evidence that the excess risk of death associated with bereavement rose with increasing number of chronic conditions (IRR for the interaction between bereaved status and number of chronic conditions 1.00, 95% CI 0.98–1.02). Accounting for the competing risk of death did not modify our main results (**Supplementary Table S26**).

## Discussion

In this population-based study, we found that spousal bereavement was associated with an excess risk of mortality and with a wide range of negative health outcomes, especially among men. The effect of bereavement reached its peak immediately after the death of the spouse and weakened over time. We observed that the illness trajectory of the deceased spouse had a substantial modifying effect on this association. Notably, the risk of non-fatal adverse health outcomes increased already in the 0-6 months before spousal loss. Our results indicate that the provision of formal and informal bereavement support should start early after, if not before, spousal loss. These findings are of particular public health and clinical relevance in the context of the Covid-19 pandemic, which will unfortunately result in a substantial number of bereaved older adults over a short period of time.

The first major conclusion of the present study is that spousal bereavement in old age seems to have a substantial causal effect on mortality and on a wide range of poor health outcomes. Our finding of a 66% increase in the risk of death from any cause during the first year of follow-up is in the upper range of previously reported estimates, both in Sweden and in other countries.^3,4^ The association between bereavement and acute cardiovascular events was similar in direction and magnitude to the findings of Carey and colleagues in the United Kingdom, despite a markedly higher rate of acute strokes among bereaved older adults in the present cohort.^18^ Not surprisingly, we found that the mortality and health disadvantage after spousal loss was greater among men than among women—with the notable exceptions of hip fractures and self-harm. Men have been shown to be more vulnerable to the acute health consequences of grief than women, most likely because of gender differences in social support, coping styles, and health behaviours around the time of bereavement.^49,50^ However, contrary to a study that investigated the relative risk of suicide after the loss of a partner in 1994–1998 in Denmark,^51^ women were found to a greater excess risk of self-harm and suicide than men after spousal loss. This was, however, mostly explained by a higher baseline incidence of self-harm among married men in the matched control group. The dramatic increase in self-harm and suicides speaks of the seriousness of the psychological distress experienced by older adults facing the loss of a partner.

The second major conclusion is that the effect of bereavement reaches its peak immediately after the death of the spouse and weakens as time elapses. The existence of a time-dependent relationship has already been suggested by previous studies, albeit with conflicting evidence.^6,15,52–54^ By using flexible parametric survival models, we were able to visualise the shape of the association between bereavement and negative health outcomes over time. Our results regarding mortality are well aligned with recent findings in Denmark, where the 2.5 times higher risk of death during the first month after spousal loss declined down to 38% six to twelve months later.^55^ Additionally, the present study shows that while the risk of acute cardiovascular events, pneumonia, and non-elective hospitalisations also increased in a transient fashion, other outcomes such as hip fractures, injuries due to self-harm, and nursing home admissions remained elevated for longer periods of time. This has important implications for public health and clinical practice, as it means that bereavement support should be offered without delay to mitigate short-term decline and then maintained over a sufficiently long period of time.

The third major conclusion is that the detrimental effects of bereavement on the surviving spouses’ health vary substantially according to the illness trajectory of the deceased. Earlier work has shown that the negative health effects of bereavement varied according to the cause of death of the deceased spouse, but most of these studies used the cause of spousal loss as a way to test for selection bias ^56^ or to separate expected from unexpected deaths.^57,58^ To our knowledge, this is the first large cohort study that has examined the impact of bereavement across different and clinically meaningful illness trajectories. We found that the excess risk of mortality was highest among bereaved older adults whose spouse died suddenly and lowest among those whose spouse died after a trajectory of prolonged dwindling (often marked by the progression of dementia or other neurodegenerative diseases). However, the latter still experience substantially higher rates of death, acute cardiovascular events, hip fractures and self-harm than their married counterparts. In that respect, our findings contrast with the conclusion by Elwert and colleagues that the spouses of people who died from Alzheimer’s disease or Parkinson’s disease remained unaffected by the widowhood effect.^59^ We also observed that bereaved older adults whose spouse died from organ failure experienced a particularly high excess risk of acute cardiovascular events and self-harm, and those whose spouse died from cancer had the highest excess risk of pneumonia after bereavement. Thus, although sudden spousal deaths expose surviving partners to especially high excess mortality, our findings suggest that longer and more predictable illness trajectories do not shield survivors from the adverse health consequences of bereavement.^60^

The fourth major conclusion is that the risk of non-fatal adverse health outcomes starts increasing already during the 6-month period *prior* to spousal loss. Our results are consistent with previous work showing substantial changes in the risk of cardiovascular events and injuries during the months before bereavement.^19,61^ In a cross-sectional analysis of the Health and Retirement Study, a nationally representative survey of Americans aged 50 years and older, researchers found that spouses nearing bereavement reported worse mobility, more impairments in instrumental activities of daily living, a greater burden of depressive symptoms and poorer working memory than continuously married individuals.^20^ The finding that health outcomes start worsening before spousal death suggests that the observed association between bereavement and ill health is partly attributable to factors that precede widowhood. Thus, any hypothesis for the causal effect of spousal bereavement on health should also explain how the experience of the surviving spouses before the death of their partners contributes to exposing them to serious adverse events.

One such hypothesis is that the effect of bereavement is mediated by enhanced inflammatory responses to psychophysiological stress and depressive symptoms.^62–64^ This exacerbated inflammation may be triggered not only by loss and grief after spousal loss, but also by ‘anticipatory grief’, namely that soon-to-be widows and widowers face substantial emotional and psychological distress before the death of their loved one actually occurs.^65^ A second hypothesis is that the “widowhood effect” stems partly from a shared health disadvantage among spouses, which could be explained both by assortative mating and by unfavourable living conditions throughout adulthood such as insufficient material resources, poor housing quality, and high-risk diet. Hence, social scientists have suggested that the excess risk of negative health events around the time of bereavement is driven either by selection mechanisms (those who become widows and widowers tend to have a lower socioeconomic position) or by the substantial reduction in material resources already before spousal loss. Although our data shows that bereaved cases have, on average, a lower level of education and income and tend to live in more socially deprived areas compared with married controls, we found no evidence of a socioeconomic gradient in the association between bereavement and mortality. We acknowledge that this may very well be specific to the study population at hand, namely older people whose main source of income was retirement pensions rather than work-related earnings, in a country where the financial toll of widowhood is automatically compensated by a specific pension scheme. Finally, a third hypothesis is that the observed health disadvantage among bereaved relatives is partly due to the caregiving burden, which is well documented among spouses of older people with advanced illness (e.g. cancer, heart failure, dementia).^66,67^ Caregiving burden is hypothesised to have a direct impact on mental health and cognition, and to affect physical health through poorer self-management of pre-existing diseases, including lower adherence to essential medicines, and postponing of routine clinical appointments.^68^ In a large cohort of 12 722 older people with chronic cardiovascular conditions in the United Kingdom, Shah and colleagues showed that during the year prior to spousal loss there was a lower uptake of basic care processes (e.g. blood pressure and HbA1c measurements) among soon-to-be widows and widowers than among their continuously married counterparts.^69^ Similarly, they found that adherence to statins, platelet antiaggregants and renin-angiotensin drugs started declining already during the months preceding spousal death. In the present study, we hypothesise that a large part of the observed increase in the risk of fall-related injuries already during the 6 months before spousal loss can be attributed to the use of sedatives, anxiolytics and hypnotics that are often prescribed to help relatives cope with the psychological stress of caring for someone at the end of life.^70,71^ This suggests that palliative care services could have an important role in detecting high-risk caregivers, providing bereavement counselling already before spousal loss, and signposting to relevant health services (including GPs and other primary care professionals) to ensure adequate management of existing conditions.

A major strength of this study is that we used routinely collected administrative and healthcare data with nationwide coverage. This eliminates the potential for non-participation bias, and thus enhances the generalizability of our findings to older people at large, at least in countries socially, culturally and economically similar to Sweden. Moreover, the use of routinely collected data mitigates the risk of recall bias. The ascertainment of the date of spousal loss was made in two separate data sources and older spouses were linked together in a deterministic rather than probabilistic manner, which provides an unbiased classification of bereaved cases and non-bereaved controls. We were able to identify and measure a wide range of demographic, socioeconomic, and health-related baseline characteristics, thus allowing us to adjust our analyses for important confounders. Matching on sex and date of birth achieved good balance of measured covariates at baseline, which suggests strong empirical equipoise and supports our claim of a causal relationship between spousal bereavement and negative health outcomes. However, the lack of information with regard to important physical (e.g. body-mass index), functional (e.g. gait speed, ADL impairments), lifestyle (e.g. diet, smoking, alcohol use) or family-related factors (e.g. level of social support) represents a potential source of residual confounding, which could threaten the validity of our effect estimates. To alleviate this concern, we calculated E-values to evaluate how susceptible the observed associations were to unmeasured confounders. This sensitivity analysis showed that unmeasured confounders would need to be of substantial magnitude to fully explain the adjusted effect estimates. For instance, the observed incidence rate ratio for all-cause mortality (1.66, 95% CI 1.53–1.80) could only be moved to the null by an unmeasured confounder associated with both spousal loss and mortality by a risk ratio of at least 2.70 above and beyond measured confounders. The existence of such confounders is, to the best of our knowledge, highly unlikely. Negative control outcomes with presumed null effect sizes were used to assess the potential for residual systemic bias, which revealed no obvious prognostic imbalance at baseline. We also devised a cohort crossover design to measure within-individual change in the risk of experiencing adverse health outcomes before and after spousal loss among bereaved cases compared with the change among married controls. This approach offers the double advantage of eliminating between-person confounding (measured or otherwise) by using study subjects as their own controls while also attenuating the risk of bias due to left truncation and natural age-related trends. Another important strength of the present study is that all prespecified sensitivity analyses and non-prespecified post-hoc analyses are reported in a clear and transparent manner and explicitly linked to the corresponding hypotheses that we intended to test.

One obvious limitation of the present study stems from its observational nature; since spousal bereavement is, by definition, a *non-randomizable* exposure, we cannot completely rule out the presence of residual confounding. Another limitation is that the categorization of end-of-life illness trajectories of the deceased spouses is both an over-simplification of the actual trajectory of functional decline that these persons followed and an imperfect proxy for the burden of caregiving that the surviving spouses experienced during the final months and weeks before spousal loss. In the self-controlled cohort crossover design, we were unable to conduct subgroup analyses by illness trajectories due to a very low number of events during each hazard period for most outcomes. Moreover, outcomes were limited to serious health events related to somatic conditions. Although we recognize that endpoints related to the development of chronic diseases, quality of life, or mental health are very important to bereaved older adults and highly relevant for clinicians and policy makers, the lack of sufficiently detailed data from primary care practitioners and the risk of detection bias (whereby bereaved individuals would, for instance, be more likely to be *reported* as having depressive symptoms and worse quality of life) prevented us from including these aspects in the present study. Finally, we were unable to investigate the long-term health consequences of spousal bereavement.

## Conclusion

In this large, population-based study, we found evidence of a substantial association spousal loss on all-cause mortality and on a wide range of negative health outcomes in old age. The hypothesis of a causal relationship was supported by the use of different epidemiological designs and a variety of statistical methods to assess and mitigate the risk of bias inherent to the observational nature of the study. Minimizing the harms of bereavement would require interventions designed to detect people at high-risk in routine practice and to offer immediate clinical and social support. Such support should be provided not only to recently bereaved spouses, but also to the spouses of seriously ill older persons with poor prognosis. Indeed, our finding that the risk of adverse health consequences increases already during the 6 months prior to spousal loss indicate that palliative care services have an important role to play in providing timely bereavement care to spouses and other family caregivers.

## Data Availability

The individual-level, pseudonymised data used in this study were provided by the National Board of Health and Welfare and by Statistics Sweden under conditions that do not permit sharing owing to privacy and confidentiality regulations. However, detailed aggregated results will be shared with the public through an interactive web application.

## Declarations

### Acknowledgments

the authors are grateful to Máté Szilcz for his help with the interactive web application, to Ben Armstrong and Antonio Gasparrini for their advices regarding statistical analyses, to Amaia Calderon Larrañaga for her constructive feedback on the manuscript, and to Marie Tournigand whose thoughtful reflections prompted this study in the first place.

## Authors contribution

LM conceptualized and designed the study, curated the data, developed the methodology, performed the statistical analysis, drafted and revised the manuscript. JW, SF, NA and KJ interpreted the data and critically revised the manuscript. KJ acquired funding and provided supervision. All authors gave approval for the final version of the manuscript and agree to be accountable for all aspects of the work.

## Ethical approval

The study was approved by the Ethical Review Board in Stockholm, Sweden (decision n°2016/1001-31/4).

## Transparency

all authors affirm that this manuscript is an honest, accurate, and transparent account of the study being reported, that no important aspects of the study have been omitted, and that any discrepancies from the study as planned have been explained.

## Availability of data and materials

the individual-level, pseudonymised data used in this study were provided by the National Board of Health and Welfare and by Statistics Sweden under conditions that do not permit sharing owing to privacy and confidentiality regulations. However, detailed aggregated results will be shared with the public through an interactive web application.

## Supplementary Tables and Figures

**Supplementary Table S1.**
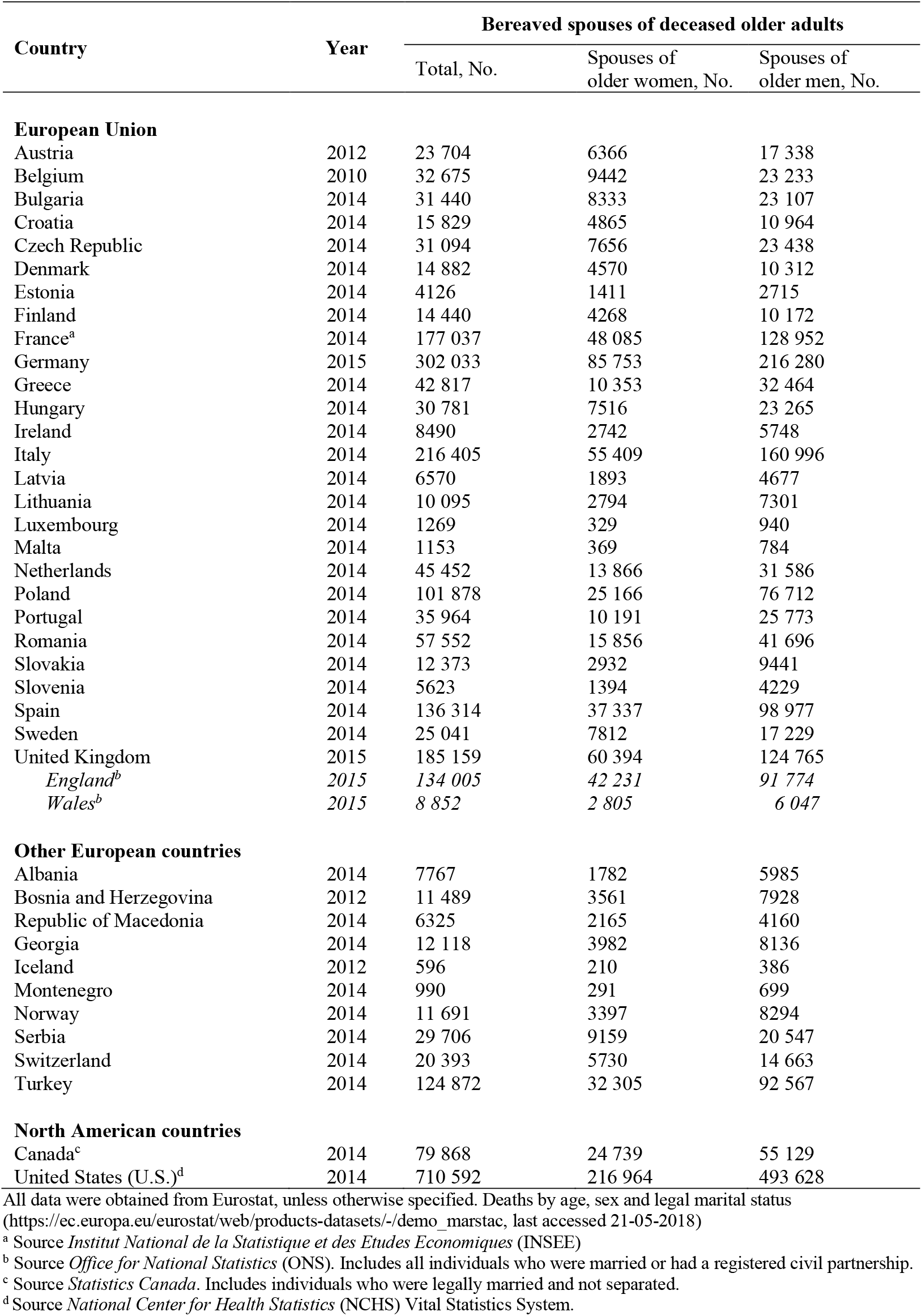
Number of individuals who experienced the loss of a spouse aged ≥65 years in selected European and Northern American countries

| Country | Year | Bereaved spouses of deceased older adults |  |  |
| --- | --- | --- | --- | --- |
|  |  | Total, No. | Spouses of older women, No. | Spouses of older men, No. |
| **European Union** |  |  |  |  |
| Austria | 2012 | 23 704 | 6366 | 17 338 |
| Belgium | 2010 | 32 675 | 9442 | 23 233 |
| Bulgaria | 2014 | 31 440 | 8333 | 23 107 |
| Croatia | 2014 | 15 829 | 4865 | 10 964 |
| Czech Republic | 2014 | 31 094 | 7656 | 23 438 |
| Denmark | 2014 | 14 882 | 4570 | 10 312 |
| Estonia | 2014 | 4126 | 1411 | 2715 |
| Finland | 2014 | 14 440 | 4268 | 10 172 |
| France<sup>a</sup> | 2014 | 177 037 | 48 085 | 128 952 |
| Germany | 2015 | 302 033 | 85 753 | 216 280 |
| Greece | 2014 | 42 817 | 10 353 | 32 464 |
| Hungary | 2014 | 30 781 | 7516 | 23 265 |
| Ireland | 2014 | 8490 | 2742 | 5748 |
| Italy | 2014 | 216 405 | 55 409 | 160 996 |
| Latvia | 2014 | 6570 | 1893 | 4677 |
| Lithuania | 2014 | 10 095 | 2794 | 7301 |
| Luxembourg | 2014 | 1269 | 329 | 940 |
| Malta | 2014 | 1153 | 369 | 784 |
| Netherlands | 2014 | 45 452 | 13 866 | 31 586 |
| Poland | 2014 | 101 878 | 25 166 | 76 712 |
| Portugal | 2014 | 35 964 | 10 191 | 25 773 |
| Romania | 2014 | 57 552 | 15 856 | 41 696 |
| Slovakia | 2014 | 12 373 | 2932 | 9441 |
| Slovenia | 2014 | 5623 | 1394 | 4229 |
| Spain | 2014 | 136 314 | 37 337 | 98 977 |
| Sweden | 2014 | 25 041 | 7812 | 17 229 |
| United Kingdom | 2015 | 185 159 | 60 394 | 124 765 |
| *England<sup>b</sup>* | 2015 | 134 005 | 42 231 | 91 774 |
| *Wales<sup>b</sup>* | 2015 | 8 852 | 2 805 | 6 047 |
| **Other European countries** |  |  |  |  |
| Albania | 2014 | 7767 | 1782 | 5985 |
| Bosnia and Herzegovina | 2012 | 11 489 | 3561 | 7928 |
| Republic of Macedonia | 2014 | 6325 | 2165 | 4160 |
| Georgia | 2014 | 12 118 | 3982 | 8136 |
| Iceland | 2012 | 596 | 210 | 386 |
| Montenegro | 2014 | 990 | 291 | 699 |
| Norway | 2014 | 11 691 | 3397 | 8294 |
| Serbia | 2014 | 29 706 | 9159 | 20 547 |
| Switzerland | 2014 | 20 393 | 5730 | 14 663 |
| Turkey | 2014 | 124 872 | 32 305 | 92 567 |
| **North American countries** |  |  |  |  |
| Canada<sup>c</sup> | 2014 | 79 868 | 24 739 | 55 129 |
| United States (U.S.)<sup>d</sup> | 2014 | 710 592 | 216 964 | 493 628 |

All data were obtained from Eurostat, unless otherwise specified. Deaths by age, sex and legal marital status (https://ec.europa.eu/eurostat/web/products-datasets/-/demo_marstac, last accessed 21-05-2018)
<sup>a</sup> Source *Institut National de la Statistique et des Etudes Economiques* (INSEE)
<sup>b</sup> Source *Office for National Statistics* (ONS). Includes all individuals who were married or had a registered civil partnership.
<sup>c</sup> Source *Statistics Canada*. Includes individuals who were legally married and not separated.
<sup>d</sup> Source *National Center for Health Statistics* (NCHS) Vital Statistics System.

**Supplementary Table S2.** Information about the data sources

| Database | Variables used in the study | Additional information |
| --- | --- | --- |
| Total Population Register (RTB) | Sex, date of birth, country of birth, municipality of residence, living arrangement, civil status, date of change in civil status, year of first and last immigration, year of last emigration | The Total Population Register (Swedish: *Registret över totalbefolkningen*) was established in 1968 by the government agency *Statistics Sweden* based on computerized records from the local population registers. It is maintained continuously and updated 5 times per week with data from the Swedish National Tax Agency. The personal identity number was first introduced in 1947. Covers all persons registered as Swedish residents, including immigrants. |
| Swedish Register of Education | Highest educational attainment | The Swedish Register of Education (Swedish: *Utbildningsregistret*) was first established in 1985. It contains data from the 1970 and 1990 population and housing censuses, as well as annual updates from Statistics Sweden from 2000 to 2014. Data in the Education Register are coded with the SUN2000 nomenclature, which is adapted to the international International Standard Classification of Education (ISCED 97). Overall, 1.7% of the study population was missing information on the level of education. |
| Longitudinal integration database for health insurance and labour market studies (LISA) | Disposable income | The LISA database (Swedish: *Longitudinell Integrationsdatabas för Sjukförsäkrings- och Arbetsmarknadsstudier*) was established in 1990 and expanded considerably in 2004. It includes all individuals aged $\geq 16$ years and registered in Sweden on 31 December of each year. Disposable income is calculated as the total income received annually minus taxes. It includes earning (e.g. salaries), business income (e.g. income from farms), sickness allowances, disability pensions, unemployment benefits, retirement and old-age pensions (National Retirement Pension Scheme), and general social support provisions. In this study, the income of each individual during the year before spousal loss was equalised by dividing the total household disposable income by the number of consumption units in the household (first adult = 1, second adult = 0.51). Only 125 persons were missing data on their income. In order to facilitate international comparisons, the disposable income per consumption unit was converted into U.S. dollars by using the Swedish Central Bank average exchange rate from 1 January to 31 December of each year (1 SEK = 0.1477 / 0.1536 USD). |
| National Cause of Death Register | Sex, age, civil status at time of death, underlying and contributing causes of death, and place of death. | The Swedish National Cause of Death Register (Swedish: *Dödsorsaksregistret*) is complete since 1952, although detailed data are available at the national level since 1911. Causes of death are coded with the International Statistical Classification of Diseases and Related Health Problems (ICD-10 version from 1997 onwards). The physician certifying death can mention up to 48 causes. Electronic certification is |
|  |  | available since 2011. Swedish residents who die abroad are included. About 1% of all deaths have no reported underlying cause of death. |
| National Patient Register | Sex, age, date of admission, elective/non-elective admission, main diagnosis, secondary diagnoses, external causes of injury, medical or surgical procedures, department of admission, provenance, destination | The National Patient Register (Swedish: *Patientregistret*) was started in 1964 and has full national coverage for inpatient care since 1987. It includes all inpatient admissions in Sweden from both public and private providers, with >99% completeness for somatic and psychiatric hospital discharges. Clinical diagnoses are coded according to the ICD classification (ICD-10 since 1997). The validity of these diagnoses has been found to be very high, with a positive predictive value >95% for acute and severe conditions. Data about non-inpatient care include specialized outpatient care visits (i.e. hospital-based outpatient care, incl. day surgery and psychiatric care) since 2001. The coverage of outpatient diagnoses is typically lower than for inpatient data but remains above 80% overall. The National Patient Register does not contain information about primary care or home care. |
| Social Services Register | Sex, age, provision of home care help (with type of ADL-help and number of hours per month), housing in a nursing home, date of enforcement | The Social Services Register (Swedish: *Socialtjänstregister*) collects individual-level information about the provision of municipal services to older persons and persons with functional impairments. It was first launched in 2007 and has been implemented in routine since 2013. The register contains data about persons who live in nursing homes and residential care homes, persons who have been granted home care help or assistance for their activities of daily living in the community, and persons who are admitted into short-term nursing facilities. The status of each individual is reported on the last day of each month (e.g. 31 October). The quality of the data reported to the Social Services Register is generally good, but some municipalities have lower-than-average reporting rates. |
| Swedish Prescribed Drug Register | Sex, age, date of prescription, date of dispensing, type of dispensing (single vs. multi-dose), total dose, prescribed daily dose (free text), DDD, ATC code, generic name, cost, characteristics of prescribers | The Swedish Prescribed Drug Register (Swedish: *Läkemedelsregistret*) was implemented in 2005. Data about all dispensed prescription drugs in Sweden are transferred on a monthly basis to the National Board of Health and Welfare. Approximately 85% of all dispensed defined daily doses (DDDs) are covered by the register. Over-the-counter medications (12% DDDs), drugs administered in hospitals (3% DDDs) are not included. Vaccines and drugs dispensed in nursing homes with their own drug storeroom are only partially covered. |

**Supplementary Table S3.** List of International Classification of Diseases, 10th revision (ICD-10) codes used to identify adverse health outcomes during follow-up

| Outcome | Source | Indicator | ICD-10 codes |
| --- | --- | --- | --- |
| All-cause mortality | Cause of Death Register | Time to death | — |
| Acute cardiovascular event (main analysis) | National Patient Register, Cause of Death Register | Time to first non-elective admission or time to death | I21, I22, I24, I26, I30, I40, I46, I61, I63, I64, I71 |
| Acute cardiovascular event (broader definition) | National Patient Register, Cause of Death Register | Time to first non-elective admission or time to death | I21, I22, I23, I24, I26, I30, I33, I35, I36, I38, I39, I40, I41, I42, I43, I46, I60, I61, I62, I63, I64, I67, I71, I72, I801, I802, I803, I808, I81, I820, I821, I822, I823, I828, I829 |
| Acute myocardial infarction | National Patient Register, Cause of Death Register | Time to first non-elective admission or time to death | I21-I22 |
| Stroke | National Patient Register, Cause of Death Register | Time to first non-elective admission or time to death | I61, I63, I64 |
| Pulmonary embolism | National Patient Register, Cause of Death Register | Time to first non-elective admission or time to death | I26 |
| Aortic aneurysm and dissection | National Patient Register, Cause of Death Register | Time to first non-elective admission or time to death | I71 |
| Pneumonia | National Patient Register, Cause of Death Register | Time to first non-elective admission or time to death | J12-J18 |
| Hip fracture | National Patient Register, Cause of Death Register | Time to first non-elective admission or time to death | S72 |
| Self-harm | National Patient Register, Cause of Death Register | Time to first non-elective admission or time to death | X60-X84, Y10-Y34 |
| Injurious fall | National Patient Register | Time to first non-elective admission | W00-W19 |
| Non-elective hospitalisation for any cause | National Patient Register | Time to first non-elective inpatient admission | — |
| Nursing home admission | Social Services Register | Time to nursing home or residential care home admission | — |

**Supplementary Table S4.**
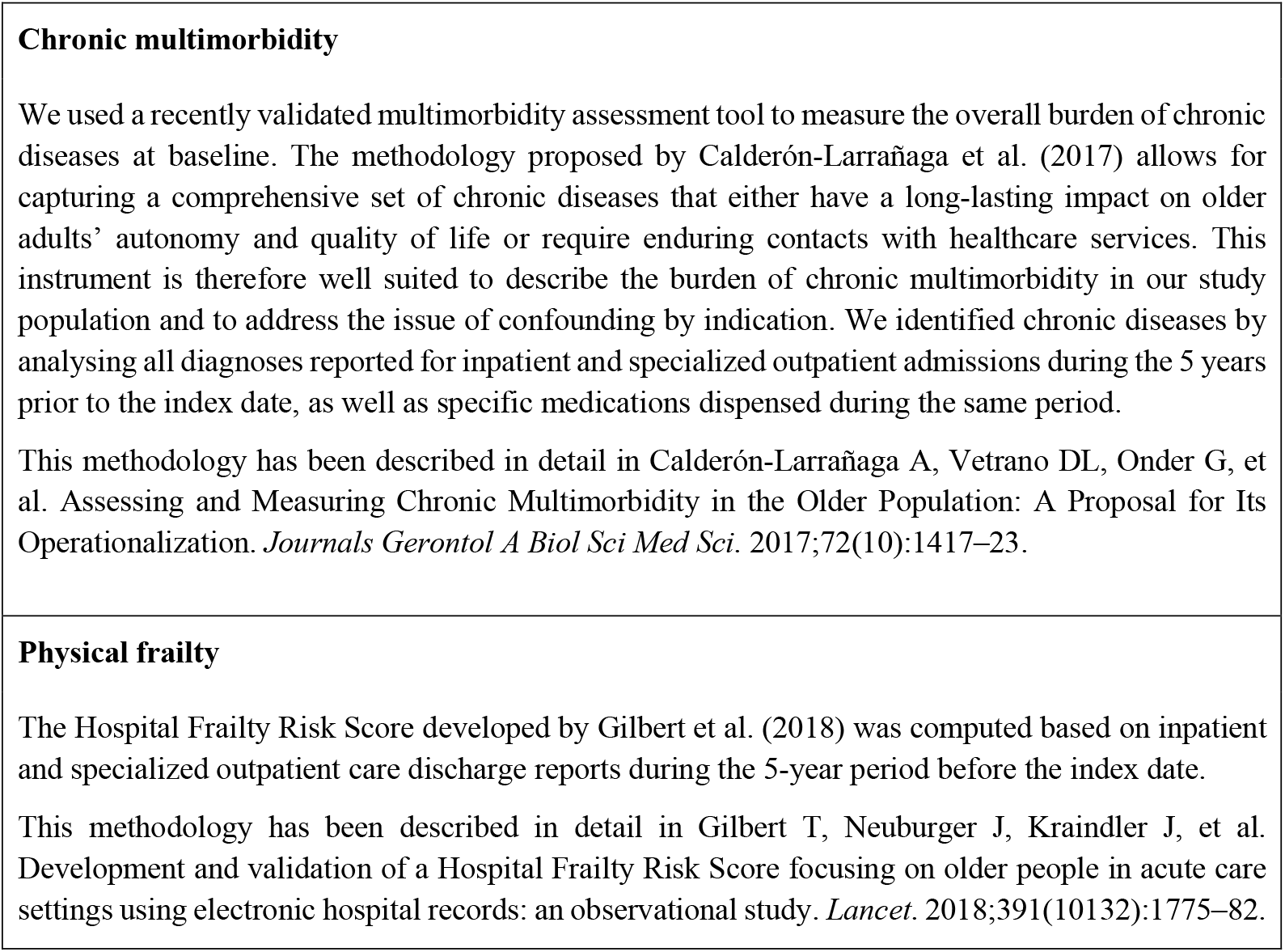
Measure of chronic multimorbidity and physical frailty at baseline

**Supplementary Table S5.** List of International Classification of Diseases, 10th revision (ICD-10) codes used to identify history of specific diseases

| **Condition** | **ICD-10 codes** |
| --- | --- |
| History of smoking-related cancer | C00, C01, C02, C03, C04, C05, C06, C07, C08, C09, C10, C11, C12, C13, C14, C15, C16, C25, C32, C33, C34, C53, C64, C65, C66, C67, C68, C09, C920, D00, D06, D019, D020, Z72, Z864, T652 |
| History of alcohol-related disease | F10, E244, G312, G621, G721, I426, K292, K70, K852, K860, X65, X45, R780, Y15, Y90, Y91 |
| History of depression and mood disorders | F30, F31, F32, F33, F34, F38, F39, F412 |
| History of other psychiatric disease | F04, F06, F07, F09, F102, F106, F107, F112, F116, F117, F122, F126, F127, F132, F136, F137, F142, F146, F147, F152, F156, F157, F162, F166, F167, F172, F176, F177, F182, F186, F187, F192, F196, F197, F20, F22, F24, F25, F28, F50, F52, F60, F61, F62, F63, F68, F70–F89, F95, F99 |

**Supplementary Table S6.** Small-area level of social deprivation

|  | Small-area level of social deprivation<br>(quartiles, from least deprived to most deprived) |  |  |  | Missing |
| --- | --- | --- | --- | --- | --- |
|  | 1 | 2 | 3 | 4 |  |
| Bereaved (cases) | 9578 (22.3%) | 9243 (21.5%) | 11 171 (26.0%) | 12 875 (30.0%) | 51 (0.1%) |
| Married (controls) | 11 262 (26.2%) | 9338 (21.8%) | 10 668 (24.9%) | 11 565 (26.9%) | 86 (0.2%) |
| Widowed (controls) | 8612 (20.1%) | 8296 (19.3%) | 10 911 (25.4%) | 15 045 (35.1%) | 54 (0.1%) |

**Supplementary Table S7.** Characteristics of bereaved cases and long-time widowed controls

|  | **Bereaved cases** | **Long-time widowed controls** |
| --- | --- | --- |
| **Total, No.** | 42 918 | 42 918 |
| **Sex** |  |  |
| Male | 13 581 (31.6%) | 13 581 (31.6%) |
| Female | 29 337 (68.4%) | 29 337 (68.4%) |
| **Age at index date** |  |  |
| Mean, SD | 78.9 (7.2) | 78.9 (7.2) |
| 65 to 74 years | 13 944 (32.5%) | 13 953 (32.5%) |
| 75 to 84 years | 19 445 (45.3%) | 19 424 (45.3%) |
| 85 to 94 years | 9320 (21.7%) | 9323 (21.7%) |
| ≥95 years | 209 (0.5%) | 218 (0.5%) |
| **Chronic diseases** |  |  |
| Mean number, SD | 2.8 (2.7) | 2.8 (2.7) |
| Atrial fibrillation | 4675 (10.9%) | 4719 (11.0%) |
| Cerebrovascular disease | 3184 (7.4%) | 3302 (7.7%) |
| Congestive heart failure | 2779 (6.5%) | 3045 (7.1%) |
| Chronic ischemic heart disease | 7810 (18.2%) | 7970 (18.6%) |
| Chronic obstructive pulmonary disease | 2449 (5.7%) | 2990 (7.0%) |
| Dementia | 1343 (3.1%) | 1230 (2.9%) |
| Diabetes | 5891 (13.7%) | 6205 (14.5%) |
| Solid cancer | 6400 (14.9%) | 6355 (14.8%) |
| **Other conditions** |  |  |
| History of smoking-related cancer | 1543 (3.6%) | 1623 (3.8%) |
| History of alcohol-related disease | 303 (0.7%) | 355 (0.8%) |
| History of depression and mood disorders | 871 (2.0%) | 915 (2.1%) |
| History of other psychiatric diseases | 698 (1.6%) | 767 (1.8%) |
| **Hospital Frailty Risk Score** |  |  |
| Mean, SD | 2.3 (3.7) | 2.3 (3.8) |
| Low risk (<5) | 36 199 (84.3) | 35 979 (83.8) |
| Intermediate risk (5-9) | 4610 (10.7) | 4647 (10.8) |
| High risk (≥10) | 2109 (4.9) | 2292 (5.4) |
| **Prescription drugs** |  |  |
| Mean number of drugs, SD | 5.7 (4.1) | 5.7 (4.2) |
| Polypharmacy (≥5 drugs) | 24 285 (56.6%) | 24 220 (56.4%) |
| Use of specific drugs: |  |  |
| Any antihypertensive | 27 515 (64.1%) | 27 559 (64.2%) |
| Statins | 12 701 (29.6%) | 12 291 (28.6%) |
| VKA and platelet antiaggregant | 17 044 (39.7%) | 17 380 (40.5%) |
| **Level of education** |  |  |
| Primary education | 16 267 (37.9%) | 17 062 (39.8%) |
| Secondary education | 19 220 (44.8%) | 18 743 (43.7%) |
| Tertiary education | 6990 (16.3%) | 6192 (14.4%) |
| Missing | 441 (1.0%) | 921 (2.1%) |
| **Disposable income, thousand US dollars** |  |  |
| Median (IQR) | 27.7 (23.4 – 34.8) | 23.5 (20.3 – 30.1) |
| **Small-area level of social deprivation** |  |  |
| 1 (least deprived areas) | 9578 (22.3%) | 8612 (20.1%) |
| 2 | 9243 (21.6%) | 8296 (19.3%) |
| 3 | 11 171 (26.0%) | 10 911 (25.4%) |
| 4 (most deprived areas) | 12 875 (30.0%) | 15 045 (35.1%) |
| Missing | 51 (0.1%) | 54 (0.1%) |

**Supplementary Figure S1.**
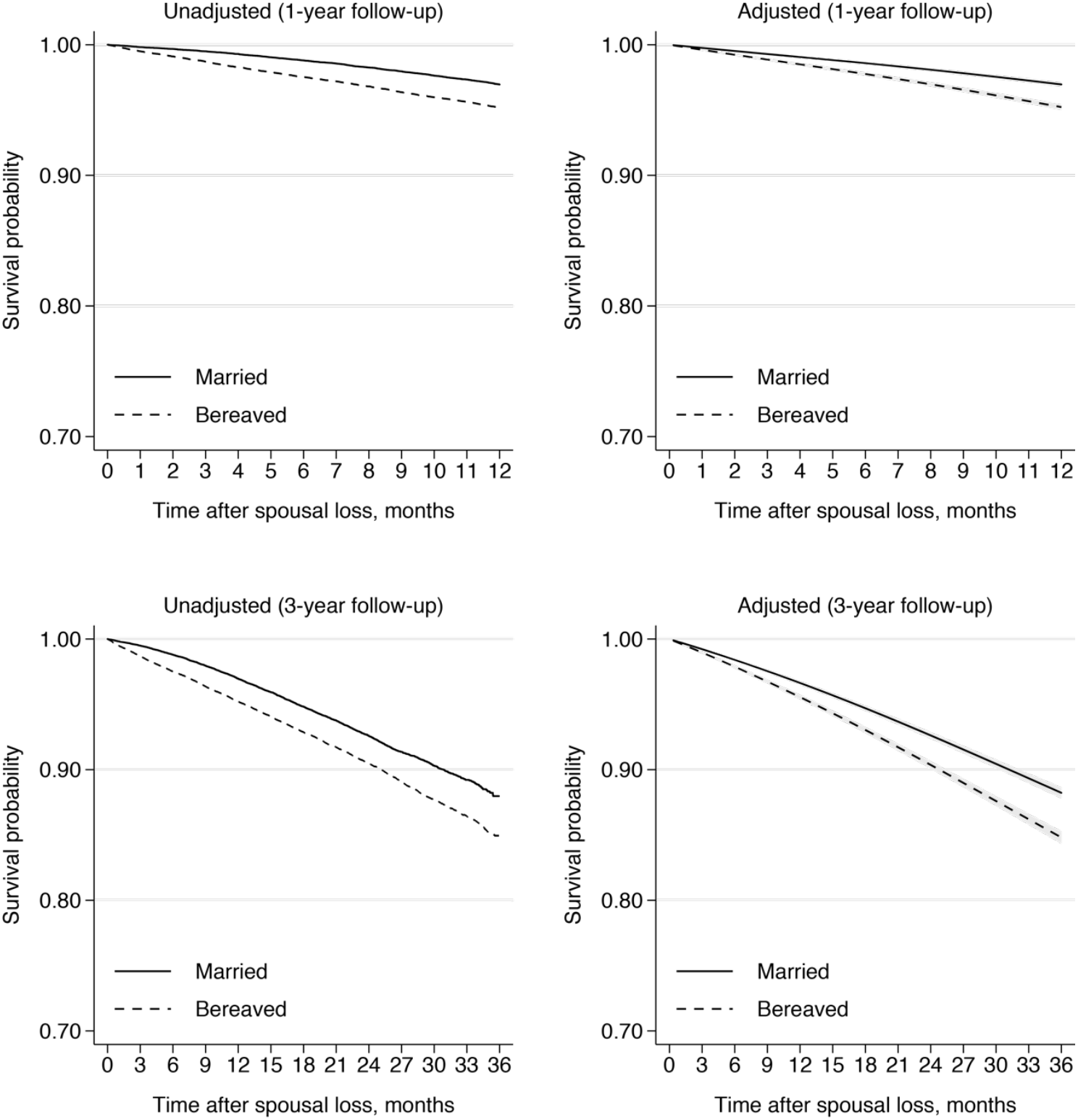
Overall survival of bereaved cases and married controls. Unadjusted curves were produced by using Kaplan-Meier estimators. Adjusted curves were produced after fitting Royston-Parmar flexible parametric survival regression models adjusted for sex, age, level of education, chronic obstructive pulmonary disease, diabetes, chronic heart failure, history of ischemic heart diseases and history of solid cancer in the previous 5 years, and number of other chronic diseases. Note that the y axis does not start at 0.00 but instead ranges from 0.70 to 1.00.

**Supplementary Table S8.** Restricted mean survival time among bereaved cases and married controls

|  | **First year of follow-up** | **Complete follow-up** |
| --- | --- | --- |
| Number of individuals | 42 918 | 42 918 |
| Restricted Mean Survival Time (95% CI), in days |  |  |
| Married controls | 360.2 (359.9 to 360.5) | 1020.9 (1019.0 to 1022.8) |
| Bereaved cases | 356.0 (355.6 to 356.5) | 1000.7 (998.5 to 1003.0) |
| Between-group difference | -4.2 (-4.7 to -3.7) | -20.2 (-23.1 to -17.2) |

**Supplementary Figure S2.**
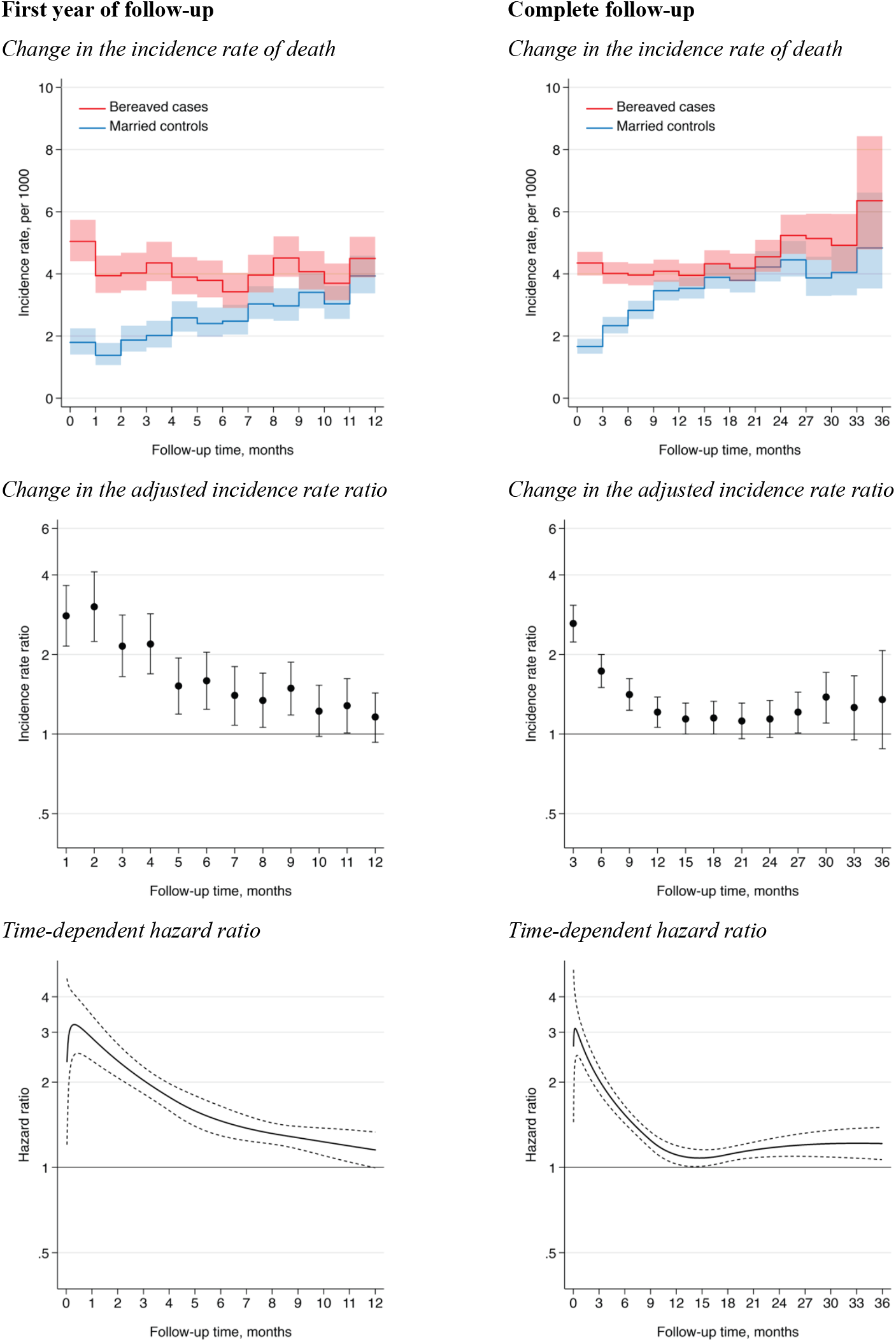
Time-varying association of spousal loss with mortality. The change in the incidence rate ratio over time was obtained from a piecewise unconditional Poisson regression model adjusted for sex, age, level of education, chronic obstructive pulmonary disease, diabetes, chronic heart failure, history of ischemic heart diseases and history of solid cancer in the previous 5 years, and number of other chronic diseases. Flexible parametric survival regression models using restricted cubic splines with 4 knots were fitted in order to plot a smooth curve of the hazard ratio for death as a function of time since spousal loss. Dotted lines indicate 95% confidence intervals.

**Supplementary Table S9.** Change in the association between spousal loss and all-cause mortality throughout follow-up

|  | Bereaved cases |  | Married controls |  | Incidence rate difference (95% CI)<sup>b</sup> | Adjusted incidence rate ratio (95% CI)<sup>c</sup> |
| --- | --- | --- | --- | --- | --- | --- |
|  | No. deaths / Time at risk | Incidence rate (95% CI)<sup>a</sup> | No. deaths / Time at risk | Incidence rate (95% CI)<sup>a</sup> |  |  |
| First year of follow-up (1-month periods) |  |  |  |  |  |  |
| Month 1 | 215 / 42 812 | 5.0 (4.4–5.7) | 77 / 42 882 | 1.8 (1.4–2.2) | 3.2 (2.4–4.0) | 2.80 (2.15–3.65) |
| Month 2 | 168 / 42 619 | 3.9 (3.4–4.6) | 59 / 42 811 | 1.4 (1.1–1.8) | 2.6 (1.9–3.3) | 3.03 (2.24–4.11) |
| Month 3 | 171 / 42 450 | 4.0 (3.5–4.7) | 80 / 42 740 | 1.9 (1.5–2.3) | 2.2 (1.4–2.9) | 2.15 (1.65–2.82) |
| Month 4 | 184 / 42 263 | 4.4 (3.8–5.0) | 86 / 42 659 | 2.0 (1.6–2.5) | 2.3 (1.6–3.1) | 2.19 (1.69–2.84) |
| Month 5 | 164 / 42 096 | 3.9 (3.3–4.5) | 110 / 42 559 | 2.6 (2.1–3.1) | 1.3 (0.5–2.1) | 1.52 (1.19–1.94) |
| Month 6 | 159 / 41 935 | 3.8 (3.2–4.4) | 102 / 42 456 | 2.4 (2.0–2.9) | 1.4 (0.6–2.1) | 1.59 (1.24–2.04) |
| Month 7 | 143 / 41 784 | 3.4 (2.9–4.0) | 105 / 42 351 | 2.5 (2.0–3.0) | 0.9 (0.2–1.7) | 1.40 (1.08–1.80) |
| Month 8 | 165 / 41 631 | 4.0 (3.4–4.6) | 128 / 42 232 | 3.0 (2.5–3.6) | 0.9 (0.1–1.7) | 1.34 (1.06–1.70) |
| Month 9 | 187 / 41 457 | 4.5 (3.9–5.2) | 125 / 42 108 | 3.0 (2.5–3.5) | 1.5 (0.7–2.4) | 1.49 (1.18–1.87) |
| Month 10 | 168 / 41 273 | 4.1 (3.5–4.7) | 143 / 41 976 | 3.4 (2.9–4.0) | 0.7 (–0.2–1.5) | 1.22 (0.98–1.53) |
| Month 11 | 152 / 41 121 | 3.7 (3.2–4.3) | 127 / 41 837 | 3.0 (2.6–3.6) | 0.7 (–0.1–1.5) | 1.28 (1.01–1.62) |
| Month 12 | 184 / 40 947 | 4.5 (3.9–5.2) | 164 / 41 698 | 3.9 (3.4–4.6) | 0.6 (–0.3–1.4) | 1.15 (0.93–1.43) |
| Complete follow-up (3-month periods) |  |  |  |  |  |  |
| Months 1 to 3 | 554 / 127 881 | 4.3 (4.0–4.7) | 216 / 128 434 | 1.7 (1.5–1.9) | 2.7 (2.2–3.1) | 2.61 (2.23–3.07) |
| Months 4 to 6 | 507 / 126 293 | 4.0 (3.7–4.4) | 298 / 127 674 | 2.3 (2.1–2.6) | 1.7 (1.2–2.1) | 1.73 (1.50–2.00) |
| Months 7 to 9 | 495 / 124 871 | 4.0 (3.6–4.3) | 358 / 126 691 | 2.8 (2.5–3.1) | 1.1 (0.7–1.6) | 1.41 (1.22–1.61) |
| Months 10 to 12 | 504 / 123 341 | 4.1 (3.7–4.5) | 434 / 125 511 | 3.5 (3.1–3.8) | 0.6 (0.1–1.1) | 1.21 (1.06–1.38) |
| Months 13 to 15 | 450 / 113 843 | 4.0 (3.6–4.3) | 410 / 116 078 | 3.5 (3.2–3.9) | 0.4 (–0.1–0.9) | 1.14 (1.00–1.31) |
| Months 16 to 18 | 423 / 97 804 | 4.3 (3.9–4.8) | 389 / 100 038 | 3.9 (3.5–4.3) | 0.4 (–0.1–1.0) | 1.15 (1.00–1.32) |
| Months 19 to 21 | 345 / 82 480 | 4.2 (3.8–4.6) | 321 / 84 614 | 3.8 (3.4–4.2) | 0.4 (–0.2–1.0) | 1.12 (0.96–1.31) |
| Months 22 to 24 | 303 / 66 602 | 4.5 (4.1–5.1) | 289 / 68 504 | 4.2 (3.8–4.7) | 0.3 (–0.4–1.0) | 1.14 (0.96–1.34) |
| Months 25 to 27 | 265 / 50 617 | 5.2 (4.6–5.9) | 233 / 52 347 | 4.5 (3.9–5.1) | 0.8 (–0.1–1.6) | 1.21 (1.01–1.44) |
| Months 28 to 30 | 186 / 36 199 | 5.1 (4.5–5.9) | 146 / 37 726 | 3.9 (3.3–4.6) | 1.3 (0.3–2.2) | 1.37 (1.10–1.71) |
| Months 31 to 33 | 111 / 22 563 | 4.9 (4.1–5.9) | 96 / 23 735 | 4.0 (3.3–4.9) | 0.9 (–0.3–2.1) | 1.26 (0.95–1.66) |
| Months 34 to 36 | 48 / 7556 | 6.4 (4.8–8.4) | 39 / 8069 | 4.8 (3.5–6.6) | 1.5 (–0.8–3.9) | 1.35 (0.88–2.07) |

<sup>a</sup> The incidence rates per 1000 person-months are identical to those reported graphically in Supplementary Figure S2 (top panel).
<sup>b</sup> Incidence rate difference is calculated as $IR_{\text{Bereaved}} - IR_{\text{Married}}$ . These represent the unadjusted absolute risk difference in mortality between bereaved cases and married controls.
<sup>c</sup> Adjusted incidence rate ratios are calculated with piecewise unconditional Poisson regression models adjusted for sex, age, level of education, chronic obstructive pulmonary disease, diabetes, chronic heart failure, history of ischemic heart diseases and history of solid cancer in the previous 5 years, and number of other chronic diseases. These incidence rate ratios are identical to those reported graphically in Supplementary Figure S2 (middle panel).

**Supplementary Figure S3.**
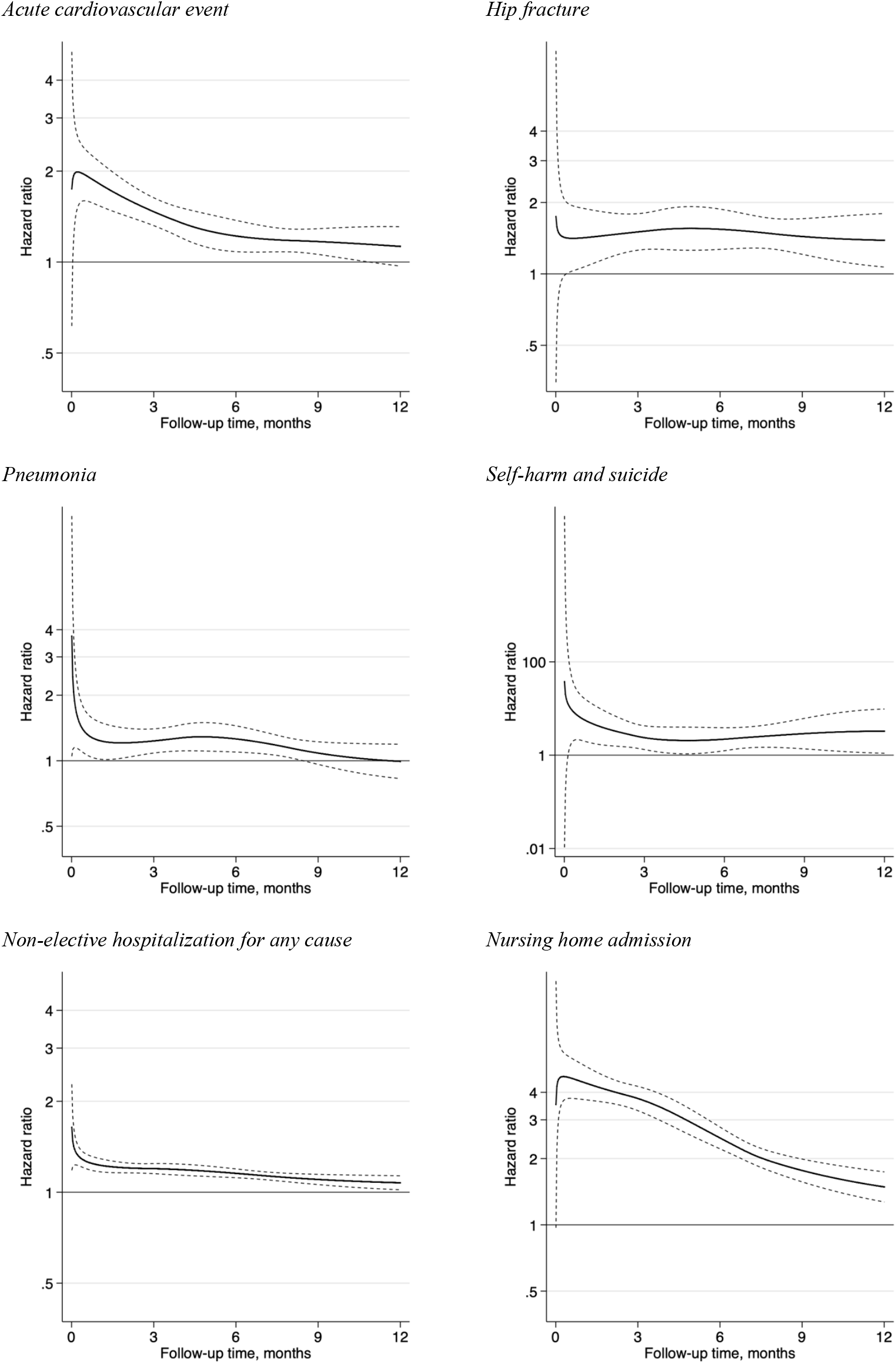
Time-varying association of spousal loss with adverse health events during the first year of follow-up.

**Supplementary Table S10.** Subgroup analysis: incidence rate ratios and 95% confidence intervals for all-cause mortality and adverse health events during the first year after spousal loss, among older men and women

|  | **Men** |  | **Women** |  |
| --- | --- | --- | --- | --- |
|  | Bereaved cases | Married controls | Bereaved cases | Married controls |
| **All-cause mortality** |  |  |  |  |
| No. of deaths | 1047 | 614 | 1013 | 692 |
| Person years of follow-up | 13 048 | 13 313 | 28 817 | 29 046 |
| Rate per 1000 person-years (95% CI) | 80.2 (75.5–85.3) | 46.1 (42.6–49.9) | 35.2 (33.1–37.4) | 23.8 (22.1–25.7) |
| Unadjusted incidence rate ratio (95% CI)<sup>a</sup> | 1.77 (1.60–1.96) |  | 1.51 (1.37–1.66) |  |
| Adjusted incidence rate ratio (95% CI)<sup>b</sup> | 1.78 (1.59–2.01) |  | 1.59 (1.42–1.79) |  |
| **Acute cardiovascular event** |  |  |  |  |
| No. of persons with the event | 830 | 610 | 995 | 785 |
| Person years of follow-up | 12782 | 13103 | 28 477 | 28 745 |
| Rate per 1000 person years (95% CI) | 64.9 (60.7–69.5) | 46.6 (43.0–50.4) | 34.9 (32.8–37.2) | 27.3 (25.5–29.3) |
| Unadjusted incidence rate ratio (95% CI)<sup>a</sup> | 1.40 (1.26–1.56) |  | 1.28 (1.17–1.41) |  |
| Adjusted incidence rate ratio (95% CI)<sup>b</sup> | 1.41 (1.25–1.58) |  | 1.29 (1.16–1.43) |  |
| **Hip fracture** |  |  |  |  |
| No. of persons with the event | 179 | 132 | 466 | 317 |
| Person years of follow-up | 12 968 | 13 249 | 28 583 | 28 876 |
| Rate per 1000 person years (95% CI) | 13.8 (11.9–16.0) | 10.0 (8.4–11.8) | 16.3 (14.9–17.9) | 11.0 (9.8–12.3) |
| Unadjusted incidence rate ratio (95% CI)<sup>a</sup> | 1.37 (1.09–1.71) |  | 1.47 (1.27–1.69) |  |
| Adjusted incidence rate ratio (95% CI)<sup>b</sup> | 1.40 (1.10–1.79) |  | 1.53 (1.31–1.77) |  |
| **Pneumonia** |  |  |  |  |
| No. of persons with the event | 556 | 480 | 599 | 520 |
| Person years of follow-up | 12 848 | 13 128 | 28 562 | 28 821 |
| Rate per 1000 person years (95% CI) | 43.3 (39.8–47.0) | 36.6 (33.4–40.0) | 21.0 (19.4–22.7) | 18.0 (16.6–19.7) |
| Unadjusted incidence rate ratio (95% CI)<sup>a</sup> | 1.17 (1.03–1.32) |  | 1.17 (1.04–1.32) |  |
| Adjusted incidence rate ratio (95% CI)<sup>b</sup> | 1.16 (1.02–1.32) |  | 1.11 (0.97–1.27) |  |
| **Self-harm (including suicide)** |  |  |  |  |
| No. of persons with the event | 32 | 16 | 57 | 14 |
| Person years of follow-up | 13 031 | 13 297 | 28 773 | 29 019 |
| Rate per 1000 person years (95% CI) | 2.5 (1.7–3.5) | 1.2 (0.7–2.0) | 2.0 (1.5–2.6) | 0.5 (0.3–0.8) |
| Unadjusted incidence rate ratio (95% CI)<sup>a</sup> | 2.00 (1.10–3.65) |  | 4.07 (2.27–7.31) |  |
| Adjusted incidence rate ratio (95% CI)<sup>b</sup> | 2.69 (1.35–5.36) |  | 4.47 (2.09–9.56) |  |

<sup>a</sup> Unadjusted conditional fixed-effect Poisson regression model with grouping on the matched sets (age), stratified by sex.
<sup>b</sup> Conditional fixed-effect Poisson regression model with grouping on the matched sets (age), stratified by sex and further adjusted on the same covariates as those reported in Table 2.

**Supplementary Table S11.**
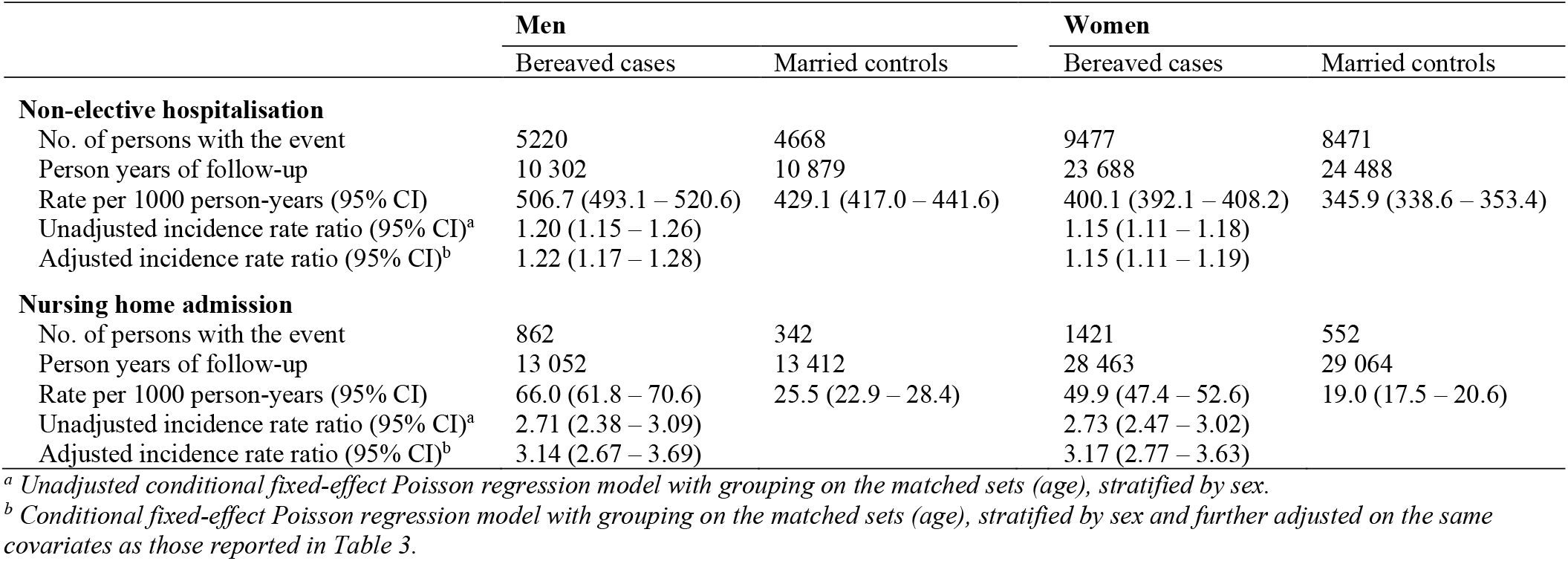
Subgroup analysis: incidence rate ratios and 95% confidence intervals for non-elective hospitalisation and nursing home admission during the first year after spousal loss, among older men and women

**Supplementary Figure S4.**
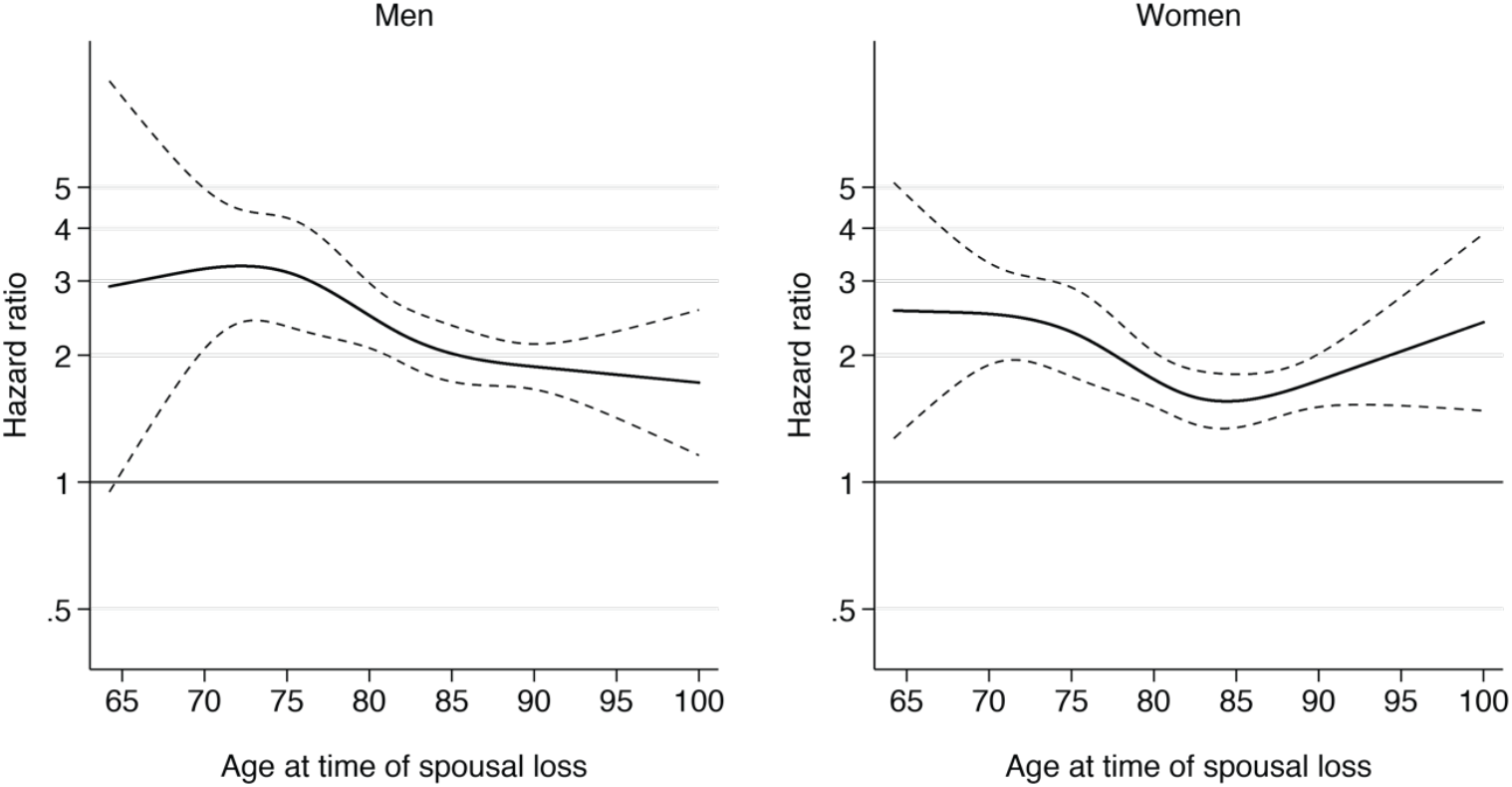
Hazard ratios for death during the first year after spousal loss according to age at baseline, among men and women. Hazard Ratios for death during the first year of follow-up according to age at time of spousal bereavement, among men and women. Solid lines indicate hazard ratios and dashed lines indicate 95% CIs. Hazard ratios were calculated by parametric survival regression analysis after adjustment for the number of chronic conditions, by using restricted cubic spline regression with 4 knots placed at the 5^th^, 35^th^, 65^th^ and 95^th^ percentiles.

**Supplementary Figure S5.**
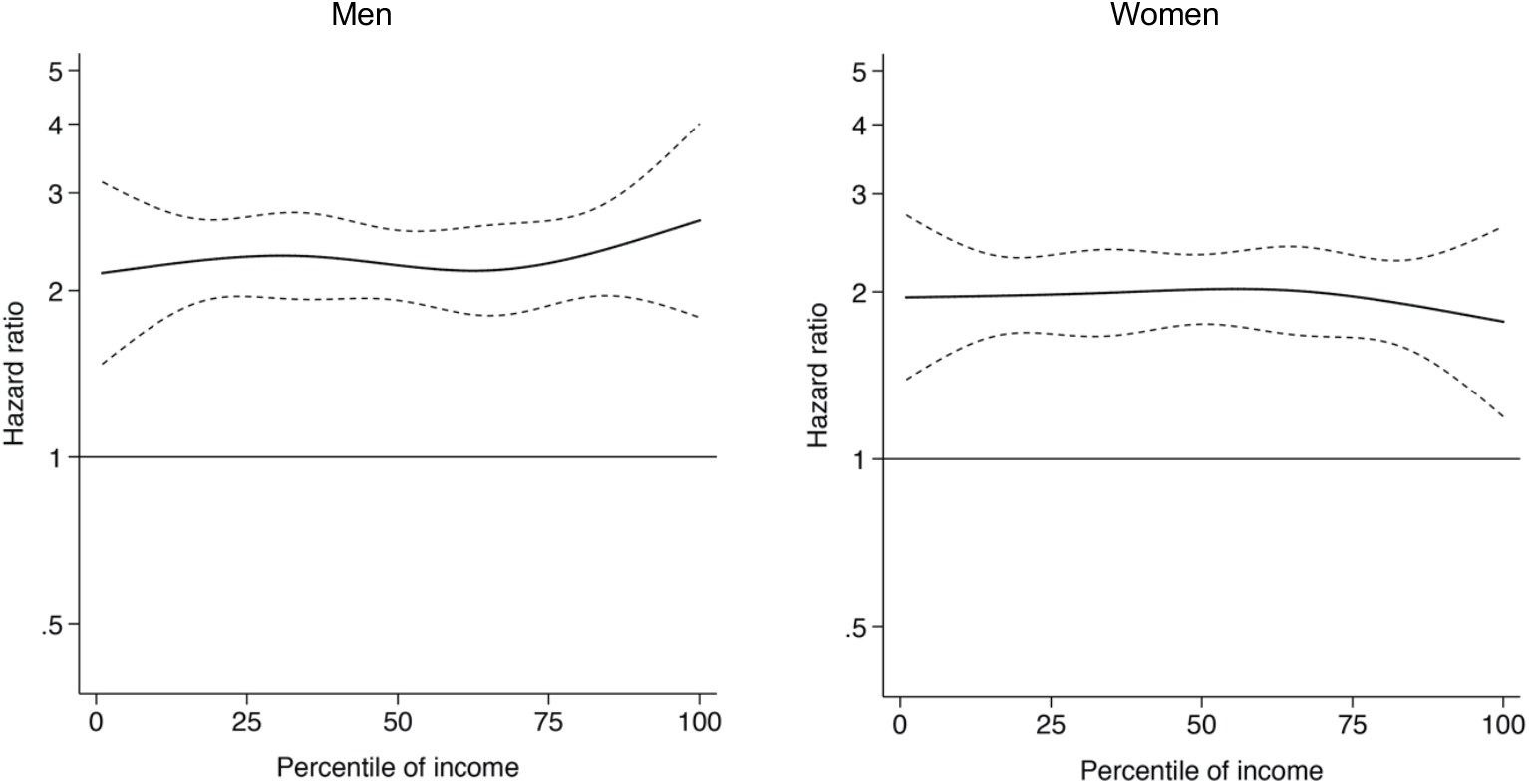
Hazard ratios for death during the first year after spousal loss according to income at baseline, among men and women. Hazard Ratios for death during the first year of follow-up according to the percentile of income at baseline, among men and women. In order to avoid reverse association, income was calculated as the disposable income per consumption unit during the year prior to spousal bereavement. Solid lines indicate hazard ratios and dashed lines indicate 95% CIs. Hazard ratios were calculated by parametric survival regression analysis after adjustment for age and for the number of chronic conditions, by using restricted cubic spline regression with 4 knots placed at the 5^th^, 35^th^, 65^th^ and 95^th^ percentiles.

**Supplementary Table S12.** Hazard ratios for death during the first year after spousal loss according to the small-area level of social deprivation

|  | Mortality rate, per 1000 (95% CI) |  | Adjusted incidence rate ratio (95% CI)<sup>a</sup> |  |
| --- | --- | --- | --- | --- |
|  | Bereaved cases | Married controls | Bereaved vs. Married | Bereaved × deprivation |
| **Quartiles of social deprivation** |  |  |  |  |
| 1 (least deprived areas) | 45.5 (41.4–50.0) | 26.3 (23.5–29.5) | 1.70 (1.46–1.98) | 1 |
| 2 | 51.3 (46.8–56.2) | 30.8 (27.4–34.6) | 1.62 (1.39–1.88) | 0.95 (0.77–1.18) |
| 3 | 48.1 (44.1–52.4) | 31.3 (28.1–34.9) | 1.61 (1.40–1.85) | 0.94 (0.77–1.15) |
| 4 (most deprived areas) | 51.6 (47.8–55.8) | 34.9 (31.6–38.5) | 1.56 (1.37–1.77) | 0.92 (0.75–1.12) |
| **Entire cohort** | 49.2 (47.1–51.4) | 30.8 (29.2–32.5) | 1.66 (1.53 – 1.80) | – |

**Supplementary Table S13.** Incidence rates and relative incidence ratios (RIR) for non-fatal adverse health outcomes before and after spousal bereavement (self-controlled cohort crossover analysis)

|  | Bereaved cases |  | Married controls |  | Bereavement × Time |
| --- | --- | --- | --- | --- | --- |
|  | No. events | Rate per 1000 person-years (95% CI) | No. events | Rate per 1000 person-years (95% CI) | Relative incidence ratio (95% CI)<sup>a</sup> |
| **Acute cardiovascular events** |  |  |  |  |  |
| Reference period | 432 | 20.3 (18.5–22.3) | 438 | 20.6 (18.7–22.6) | 1 |
| 365 to 184 days before spousal loss | 461 | 21.7 (19.8–23.7) | 435 | 20.4 (18.6–22.5) | 1.08 (0.89–1.30) |
| 183 to 1 days before spousal loss | 577 | 27.2 (25.0–29.5) | 516 | 24.3 (22.3–26.5) | 1.14 (0.95–1.36) |
| 1 to 183 days after spousal loss | 761 | 36.1 (33.6–38.8) | 591 | 27.8 (25.6–30.1) | 1.34 (1.12–1.60) |
| 184 to 365 days after spousal loss | 764 | 37.3 (34.7–40.0) | 691 | 33.1 (30.7–35.6) | 1.20 (1.00–1.43) |
| **Hip fracture** |  |  |  |  |  |
| Reference period | 183 | 8.6 (7.4–9.9) | 151 | 7.1 (6.0–8.3) | 1 |
| 365 to 184 days before spousal loss | 164 | 7.7 (6.6–9.0) | 169 | 7.9 (6.8–9.2) | 0.79 (0.58–1.08) |
| 183 to 1 days before spousal loss | 274 | 12.8 (11.4–14.5) | 200 | 9.4 (8.2–10.8) | 1.13 (0.85–1.52) |
| 1 to 183 days after spousal loss | 340 | 16.1 (14.4–17.9) | 223 | 10.5 (9.2–12.0) | 1.27 (0.96–1.70) |
| 184 to 365 days after spousal loss | 315 | 15.2 (13.6–17.0) | 236 | 11.2 (9.9–12.8) | 1.17 (0.87–1.56) |
| **Pneumonia** |  |  |  |  |  |
| Reference period | 277 | 13.0 (11.5–14.6) | 291 | 13.7 (12.2–15.3) | 1 |
| 365 to 184 days before spousal loss | 334 | 15.7 (14.1–17.5) | 299 | 14.0 (12.5–15.7) | 1.18 (0.94–1.48) |
| 183 to 1 days before spousal loss | 389 | 18.3 (16.5–20.2) | 327 | 15.3 (13.8–17.1) | 1.25 (1.00–1.57) |
| 1 to 183 days after spousal loss | 575 | 27.2 (25.1–29.6) | 473 | 22.1 (20.2–24.2) | 1.31 (1.06–1.62) |
| 184 to 365 days after spousal loss | 552 | 26.6 (24.5–29.0) | 515 | 24.6 (22.5–26.8) | 1.21 (0.97–1.49) |
| **Self-harm, excluding suicide** |  |  |  |  |  |
| Reference period | 13 | 0.6 (0.4–1.0) | 15 | 0.7 (0.4–1.2) | 1 |
| 365 to 184 days before spousal loss | 14 | 0.7 (0.4–1.1) | 12 | 0.6 (0.3–1.0) | 1.39 (0.48–4.03) |
| 183 to 1 days before spousal loss | 28 | 1.3 (0.9–1.9) | 11 | 0.5 (0.3–0.9) | 3.08 (1.09–8.71) |
| 1 to 183 days after spousal loss | 41 | 1.9 (1.4–2.6) | 17 | 0.8 (0.5–1.3) | 2.95 (1.12–7.78) |
| 184 to 365 days after spousal loss | 21 | 1.0 (0.7–1.6) | 11 | 0.5 (0.3–0.9) | 2.33 (0.78–6.97) |

<sup>a</sup> These estimates are identical to those reported graphically in Figure 3. Relative incidence ratios were calculated by running conditional fixed-effect Poisson regression with a bereavement-by-time interaction term. Estimates can be interpreted as the relative increase in the risk of experiencing adverse health outcomes among bereaved cases compared with the increase among married controls.

**Supplementary Table S14.** Sensitivity analysis: Incidence rates and relative incidence ratios (RIR) for injurious falls before and after spousal bereavement (self-controlled cohort crossover analysis)

|  | Bereaved cases |  | Married controls |  | Bereavement × Time |
| --- | --- | --- | --- | --- | --- |
|  | No. events | Rate per 1000 person-years (95% CI) | No. events | Rate per 1000 person-years (95% CI) | Relative incidence ratio (95% CI)<sup>b</sup> |
| **Injurious fall<sup>a</sup>** |  |  |  |  |  |
| Reference period | 966 | 45.7 (42.9 to 48.7) | 924 | 43.7 (40.9 to 46.6) | 1 |
| 365 to 184 days before spousal loss | 1040 | 49.2 (46.3 to 52.3) | 921 | 43.5 (40.8 to 46.4) | 1.08 (0.95 to 1.23) |
| 183 to 1 days before spousal loss | 1265 | 60.0 (56.8 to 63.4) | 1045 | 49.5 (46.6 to 52.6) | 1.16 (1.03 to 1.32) |
| 1 to 183 days after spousal loss | 1601 | 76.7 (73.1 to 80.6) | 1179 | 55.7 (52.6 to 59.0) | 1.33 (1.18 to 1.50) |
| 184 to 365 days after spousal loss | 1525 | 75.1 (71.4 to 79.0) | 1273 | 61.4 (58.1 to 64.8) | 1.21 (1.07 to 1.36) |

<sup>a</sup> Unplanned hospitalisation for fall (ICD-10 codes W00-W19).
<sup>b</sup> Relative incidence ratios (RIR) were calculated by running conditional fixed-effect Poisson regression with a bereavement-by-time interaction term. These estimates can be interpreted as the relative increase in the risk of experiencing adverse health outcomes among bereaved cases compared with the increase among married controls.

**Supplementary Table S15.**
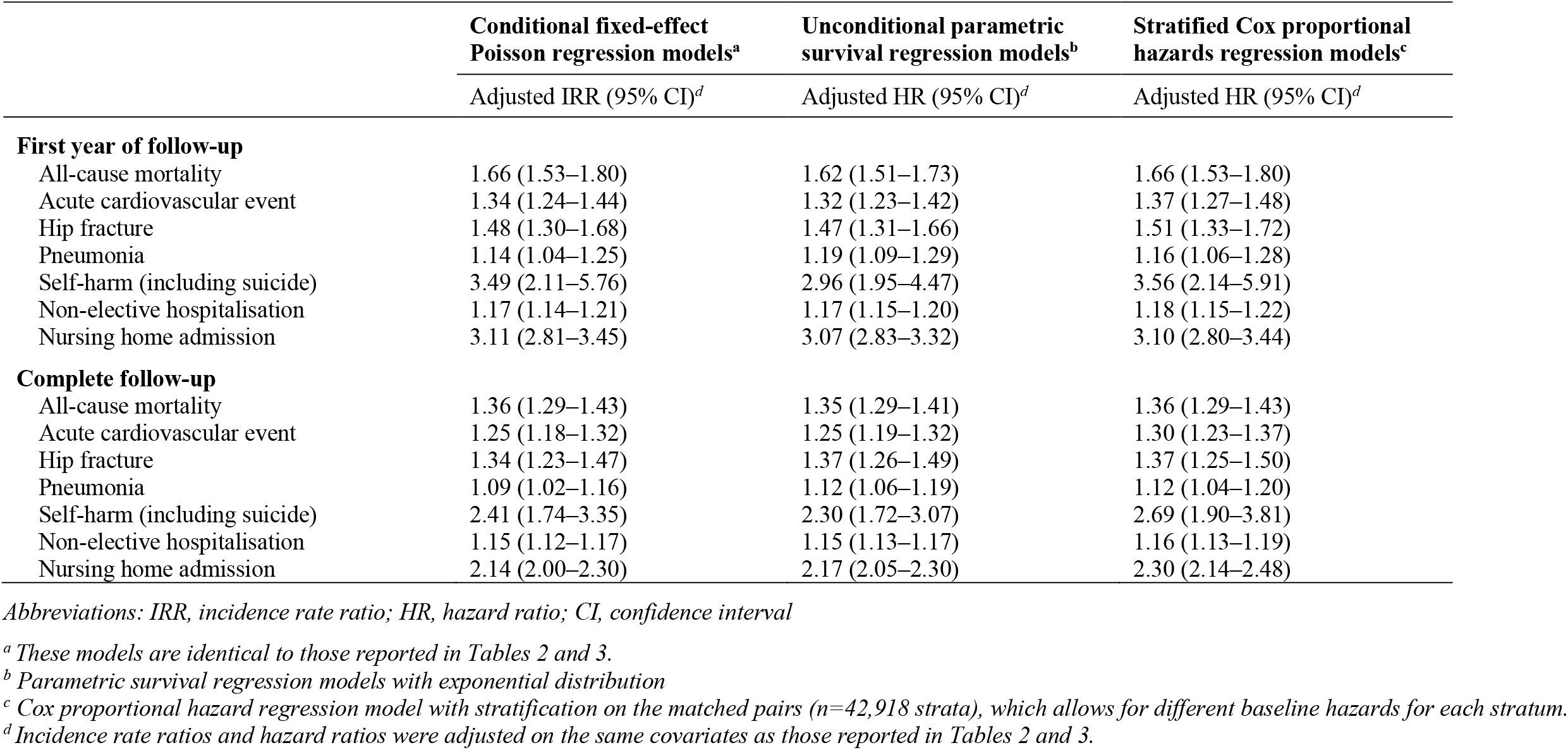
Sensitivity analysis: comparison of conditional fixed-effect Poisson regression with alternative analyses

|  | **Conditional fixed-effect**<br>**Poisson regression models<sup>a</sup>** | **Unconditional parametric**<br>**survival regression models<sup>b</sup>** | **Stratified Cox proportional**<br>**hazards regression models<sup>c</sup>** |
| --- | --- | --- | --- |
|  | Adjusted IRR (95% CI)<sup>d</sup> | Adjusted HR (95% CI)<sup>d</sup> | Adjusted HR (95% CI)<sup>d</sup> |
| **First year of follow-up** |  |  |  |
| All-cause mortality | 1.66 (1.53–1.80) | 1.62 (1.51–1.73) | 1.66 (1.53–1.80) |
| Acute cardiovascular event | 1.34 (1.24–1.44) | 1.32 (1.23–1.42) | 1.37 (1.27–1.48) |
| Hip fracture | 1.48 (1.30–1.68) | 1.47 (1.31–1.66) | 1.51 (1.33–1.72) |
| Pneumonia | 1.14 (1.04–1.25) | 1.19 (1.09–1.29) | 1.16 (1.06–1.28) |
| Self-harm (including suicide) | 3.49 (2.11–5.76) | 2.96 (1.95–4.47) | 3.56 (2.14–5.91) |
| Non-elective hospitalisation | 1.17 (1.14–1.21) | 1.17 (1.15–1.20) | 1.18 (1.15–1.22) |
| Nursing home admission | 3.11 (2.81–3.45) | 3.07 (2.83–3.32) | 3.10 (2.80–3.44) |
| **Complete follow-up** |  |  |  |
| All-cause mortality | 1.36 (1.29–1.43) | 1.35 (1.29–1.41) | 1.36 (1.29–1.43) |
| Acute cardiovascular event | 1.25 (1.18–1.32) | 1.25 (1.19–1.32) | 1.30 (1.23–1.37) |
| Hip fracture | 1.34 (1.23–1.47) | 1.37 (1.26–1.49) | 1.37 (1.25–1.50) |
| Pneumonia | 1.09 (1.02–1.16) | 1.12 (1.06–1.19) | 1.12 (1.04–1.20) |
| Self-harm (including suicide) | 2.41 (1.74–3.35) | 2.30 (1.72–3.07) | 2.69 (1.90–3.81) |
| Non-elective hospitalisation | 1.15 (1.12–1.17) | 1.15 (1.13–1.17) | 1.16 (1.13–1.19) |
| Nursing home admission | 2.14 (2.00–2.30) | 2.17 (2.05–2.30) | 2.30 (2.14–2.48) |

Abbreviations: IRR, incidence rate ratio; HR, hazard ratio; CI, confidence interval
<sup>a</sup> These models are identical to those reported in Tables 2 and 3.
<sup>b</sup> Parametric survival regression models with exponential distribution
<sup>c</sup> Cox proportional hazard regression model with stratification on the matched pairs ( $n=42,918$ strata), which allows for different baseline hazards for each stratum.
<sup>d</sup> Incidence rate ratios and hazard ratios were adjusted on the same covariates as those reported in Tables 2 and 3.

**Supplementary Table S16.** Sensitivity analysis: incidence rate ratios and 95% confidence intervals for all-cause mortality and adverse health outcomes after spousal loss among bereaved cases compared with long-time widowed controls

|  | First year of follow-up |  | Complete follow-up |  |
| --- | --- | --- | --- | --- |
|  | Bereaved cases | Long-time widowed controls | Bereaved cases | Long-time widowed controls |
| **All-cause mortality** |  |  |  |  |
| No. of deaths | 2060 | 1516 | 4191 | 3631 |
| Person years of follow-up | 41 866 | 42 240 | 81 671 | 82 901 |
| Rate per 1000 person-years (95% CI) | 49.2 (47.1–51.4) | 35.9 (34.1–37.7) | 51.3 (49.8–52.9) | 43.8 (42.4–45.2) |
| Unadjusted incidence rate ratio (95% CI)<sup>a</sup> | 1.40 (1.31–1.50) |  | 1.20 (1.15–1.26) |  |
| Adjusted incidence rate ratio (95% CI)<sup>b</sup> | 1.45 (1.34–1.57) |  | 1.24 (1.18–1.30) |  |
| **Acute cardiovascular event** |  |  |  |  |
| No. of persons with the event | 1825 | 1582 | 3396 | 3108 |
| Person years of follow-up | 41 259 | 41 654 | 79 481 | 80 743 |
| Rate per 1000 person years (95% CI) | 44.2 (42.2–46.3) | 38.0 (36.2–39.9) | 42.7 (41.3–44.2) | 38.5 (37.2–39.9) |
| Unadjusted incidence rate ratio (95% CI)<sup>a</sup> | 1.15 (1.08–1.23) |  | 1.09 (1.03–1.14) |  |
| Adjusted incidence rate ratio (95% CI)<sup>b</sup> | 1.17 (1.09–1.26) |  | 1.11 (1.06–1.17) |  |
| **Hip fracture** |  |  |  |  |
| No. of persons with the event | 645 | 535 | 1241 | 1141 |
| Person years of follow-up | 41 551 | 41 983 | 80 520 | 81 916 |
| Rate per 1000 person years (95% CI) | 15.5 (14.4–16.8) | 12.7 (11.7–13.9) | 15.4 (14.6–16.3) | 13.9 (13.1–14.8) |
| Unadjusted incidence rate ratio (95% CI)<sup>a</sup> | 1.20 (1.07–1.35) |  | 1.08 (0.99–1.17) |  |
| Adjusted incidence rate ratio (95% CI)<sup>b</sup> | 1.22 (1.08–1.38) |  | 1.07 (0.99–1.16) |  |
| **Pneumonia** |  |  |  |  |
| No. of persons with the event | 1155 | 1059 | 2153 | 2142 |
| Person years of follow-up | 41 410 | 41 815 | 80 086 | 81 367 |
| Rate per 1000 person years (95% CI) | 27.9 (26.3–29.5) | 25.3 (23.8–26.9) | 26.9 (25.8–28.0) | 26.3 (25.2–27.5) |
| Unadjusted incidence rate ratio (95% CI)<sup>a</sup> | 1.09 (1.00–1.19) |  | 1.01 (0.95–1.07) |  |
| Adjusted incidence rate ratio (95% CI)<sup>b</sup> | 1.10 (1.01–1.21) |  | 1.02 (0.96–1.09) |  |
| **Self-harm** |  |  |  |  |
| No. of persons with the event | 89 | 40 | 150 | 87 |
| Person years of follow-up | 41 803 | 42 196 | 81 560 | 82 832 |
| Rate per 1000 person years (95% CI) | 2.1 (1.7–2.6) | 0.9 (0.7–1.3) | 1.8 (1.6–2.2) | 1.1 (0.9–1.3) |
| Unadjusted incidence rate ratio (95% CI)<sup>a</sup> | 2.23 (1.53–3.23) |  | 1.71 (1.32–2.23) |  |
| Adjusted incidence rate ratio (95% CI)<sup>b</sup> | 2.60 (1.68–4.01) |  | 1.89 (1.41–2.53) |  |

**Supplementary Table S17.** Sensitivity analysis: incidence rate ratios and 95% confidence intervals for non-elective hospitalisation and nursing home admission after spousal loss among bereaved cases compared with long-time widowed controls

|  | First year of follow-up |  | Complete follow-up |  |
| --- | --- | --- | --- | --- |
|  | Bereaved cases | Long-time widowed controls | Bereaved cases | Long-time widowed controls |
| **Non-elective hospitalisation** |  |  |  |  |
| No. of persons with the event | 14 697 | 13 746 | 21 650 | 20 915 |
| Person years of follow-up | 33 989 | 34 982 | 56 823 | 59 081 |
| Rate per 1000 person years (95% CI) | 432.4 (425.5–439.5) | 392.9 (386.4–399.6) | 381.0 (375.9–385.1) | 354.0 (349.2–358.8) |
| Unadjusted incidence rate ratio (95% CI)<sup>a</sup> | 1.11 (1.08–1.13) |  | 1.07 (1.05–1.10) |  |
| Adjusted incidence rate ratio (95% CI)<sup>b</sup> | 1.14 (1.10–1.17) |  | 1.09 (1.07–1.12) |  |
| **Nursing home admission** |  |  |  |  |
| No. of persons with the event | 2289 | 1277 | 3465 | 2632 |
| Person years of follow-up | 40 658 | 41 701 | 78 236 | 80 757 |
| Rate per 1000 person years (95% CI) | 56.3 (54.0–58.6) | 30.6 (29.0–32.3) | 44.3 (42.8–45.8) | 32.6 (31.4–33.9) |
| Unadjusted incidence rate ratio (95% CI)<sup>a</sup> | 1.88 (1.75–2.02) |  | 1.40 (1.33–1.48) |  |
| Adjusted incidence rate ratio (95% CI)<sup>b</sup> | 1.94 (1.78–2.11) |  | 1.43 (1.35–1.52) |  |

<sup>a</sup> Unadjusted conditional fixed-effect Poisson regression model with grouping on the matched sets.
<sup>b</sup> Conditional fixed-effect Poisson regression model with grouping on the matched sets (sex and age), and further adjusted on the same covariates as those reported in Table 3.

**Supplementary Table S18.**
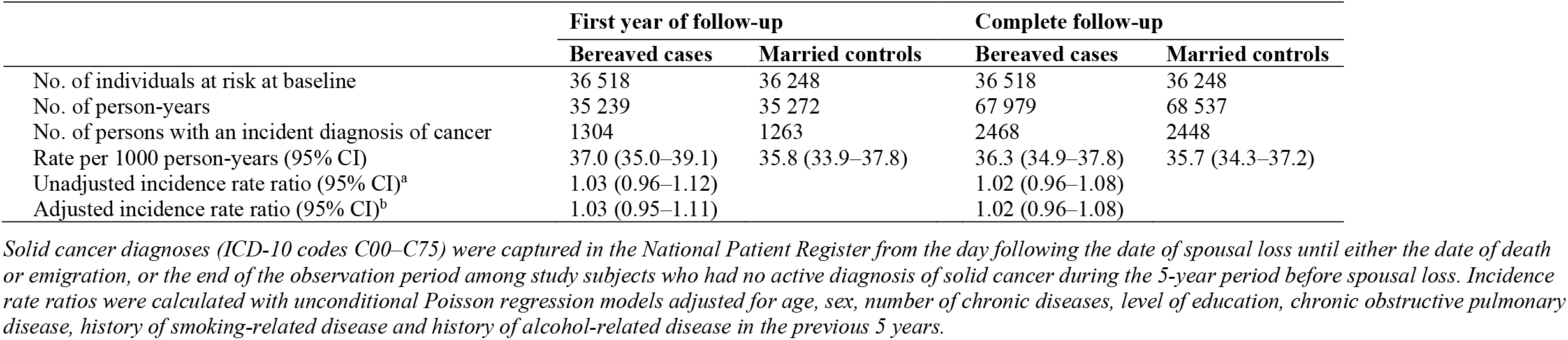
Negative control outcome: risk of being diagnosed with a solid tumour after bereavement among cancer-free individuals at baseline

|  | First year of follow-up |  | Complete follow-up |  |
| --- | --- | --- | --- | --- |
|  | Bereaved cases | Married controls | Bereaved cases | Married controls |
| No. of individuals at risk at baseline | 36 518 | 36 248 | 36 518 | 36 248 |
| No. of person-years | 35 239 | 35 272 | 67 979 | 68 537 |
| No. of persons with an incident diagnosis of cancer | 1304 | 1263 | 2468 | 2448 |
| Rate per 1000 person-years (95% CI) | 37.0 (35.0–39.1) | 35.8 (33.9–37.8) | 36.3 (34.9–37.8) | 35.7 (34.3–37.2) |
| Unadjusted incidence rate ratio (95% CI)<sup>a</sup> | 1.03 (0.96–1.12) |  | 1.02 (0.96–1.08) |  |
| Adjusted incidence rate ratio (95% CI)<sup>b</sup> | 1.03 (0.95–1.11) |  | 1.02 (0.96–1.08) |  |

*Solid cancer diagnoses (ICD-10 codes C00–C75) were captured in the National Patient Register from the day following the date of spousal loss until either the date of death or emigration, or the end of the observation period among study subjects who had no active diagnosis of solid cancer during the 5-year period before spousal loss. Incidence rate ratios were calculated with unconditional Poisson regression models adjusted for age, sex, number of chronic diseases, level of education, chronic obstructive pulmonary disease, history of smoking-related disease and history of alcohol-related disease in the previous 5 years.*

**Supplementary Figure S6.**
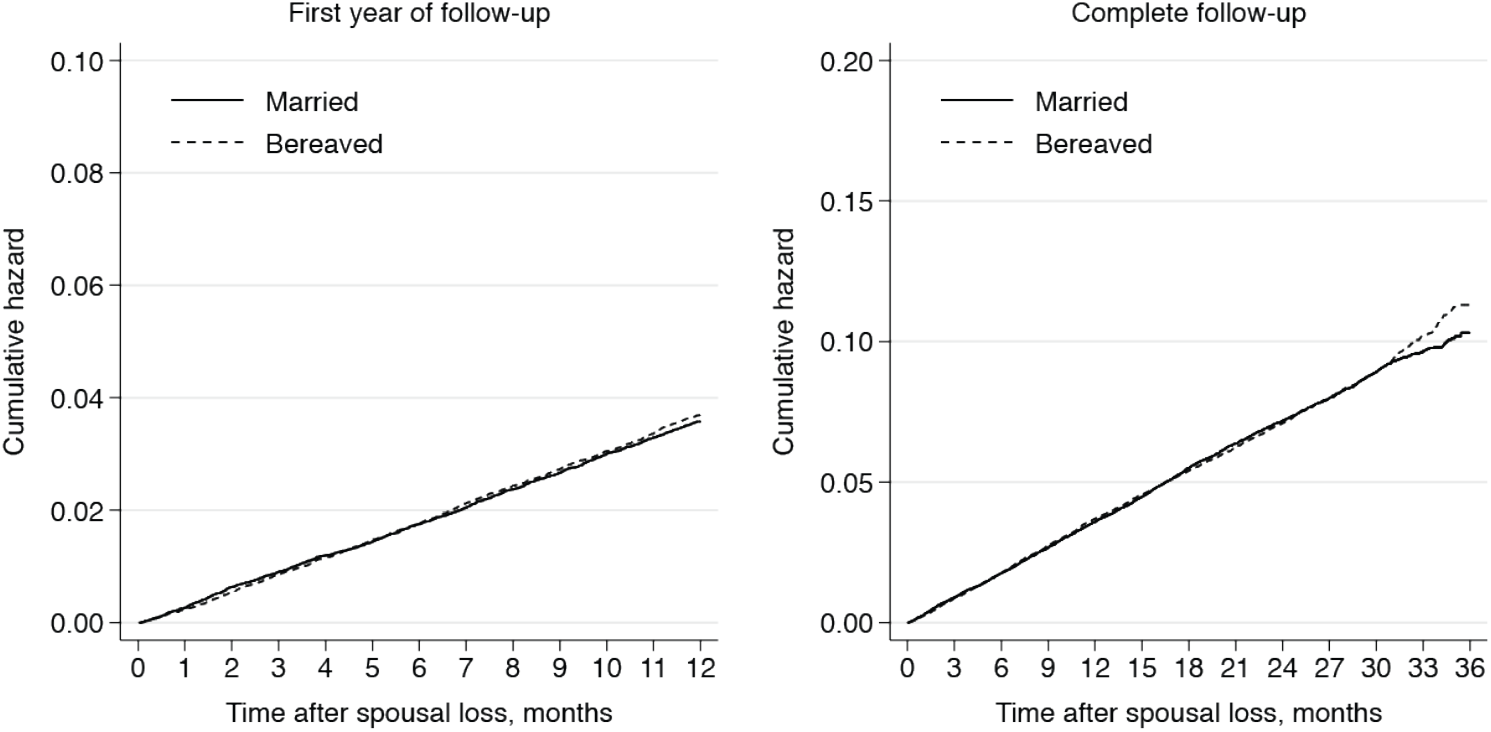
Negative control outcome: cumulative hazard of solid tumour diagnosis after bereavement among cancer-free individuals at baseline.

**Supplementary Table S19.** Negative control outcomes: hallux valgus, Parkinson’s disease, cataract

|  | First year of follow-up |  | Complete follow-up |  |
| --- | --- | --- | --- | --- |
|  | Bereaved cases | Married controls | Bereaved cases | Married controls |
| **Hallux valgus** |  |  |  |  |
| No. of individuals free of hallux valgus at baseline | 42 552 | 42 468 | 42 552 | 42 468 |
| No. of person-years | 41 441 | 41 849 | 80 785 | 82 222 |
| No. of persons with incident diagnosis of hallux valgus | 71 | 66 | 142 | 144 |
| Rate per 1000 person-years (95% CI) | 1.71 (1.36–2.16) | 1.58 (1.24–2.01) | 1.76 (1.49–2.07) | 1.75 (1.76–2.06) |
| Unadjusted incidence rate ratio (95% CI) | 1.09 (0.78–1.52) |  | 1.00 (0.80–1.27) |  |
| Adjusted incidence rate ratio (95% CI)<sup>b</sup> | 1.08 (0.77–1.51) |  | 1.00 (0.79–1.26) |  |
| **Parkinson's disease** |  |  |  |  |
| No. of individuals free of Parkinson's disease at baseline | 41 601 | 41 551 | 41 601 | 41 551 |
| No. of person-years | 40 558 | 40 981 | 79 154 | 80 628 |
| No. of persons with incident diagnosis of Parkinson's disease | 27 | 40 | 65 | 90 |
| Rate per 1000 person-years (95% CI) | 0.67 (0.46–0.97) | 0.98 (0.72–1.33) | 0.82 (0.64–1.05) | 1.12 (0.91–1.37) |
| Unadjusted incidence rate ratio (95% CI) | 0.68 (0.42–1.11) |  | 0.74 (0.53–1.01) |  |
| Adjusted incidence rate ratio (95% CI)<sup>c</sup> | 0.69 (0.42–1.12) |  | 0.74 (0.54–1.02) |  |
| **Cataract** |  |  |  |  |
| No. of individuals without history of cataract at baseline | 37 851 | 37 713 | 37 851 | 37 713 |
| No. of person-years | 35 249 | 35 528 | 66 115 | 67 271 |
| No. of persons with incident diagnosis of cataract | 3114 | 3090 | 5078 | 5057 |
| Rate per 1000 person-years (95% CI) | 88.3 (85.3–91.5) | 87.0 (84.0–90.1) | 76.8 (74.7–78.9) | 75.2 (73.1–77.3) |
| Unadjusted incidence rate ratio (95% CI) | 1.02 (0.97–1.07) |  | 1.02 (0.98–1.06) |  |
| Adjusted incidence rate ratio (95% CI)<sup>c</sup> | 1.00 (0.96–1.06) |  | 1.01 (0.97–1.05) |  |

Negative control outcomes were captured in the National Patient Register from the day following the date of spousal loss until either the date of death or emigration, or the end of the observation period. Hallux valgus: ICD-10 code M20.1; Parkinson's disease: ICD-10 code G20 ; cataract: ICD-10 code H25.
<sup>a</sup> Unconditional Poisson regression model adjusted for age, sex, history of osteoarthritis in the previous 5 years and number of other chronic diseases.
<sup>b</sup> Unconditional Poisson regression model adjusted for age, sex, number of chronic diseases, and level of education.
<sup>c</sup> Unconditional Poisson regression model adjusted for age, sex, history of diabetes or hypertension in the previous 5 years, use of antihypertensives at baseline, number of other chronic diseases and level of education.

**Supplementary Table S20.** Sensitivity analysis: E-values

|  | **First year of follow-up** |  | **Complete follow-up** |  |
| --- | --- | --- | --- | --- |
|  | Observed IRR (95% CI)<sup>a</sup> | E-value<sup>b</sup> | Observed IRR (95% CI)<sup>a</sup> | E-value<sup>b</sup> |
| **Conditional fixed-effect Poisson regression** |  |  |  |  |
| All-cause mortality | 1.66 (1.53–1.80) | 2.70 (2.43) | 1.36 (1.29–1.43) | 2.05 (1.89) |
| Acute cardiovascular event | 1.34 (1.24–1.44) | 2.01 (1.78) | 1.25 (1.18–1.32) | 1.80 (1.64) |
| Hip fracture | 1.48 (1.30–1.68) | 2.32 (1.93) | 1.34 (1.23–1.47) | 2.02 (1.76) |
| Pneumonia | 1.14 (1.04–1.25) | 1.54 (1.24) | 1.09 (1.02–1.16) | 1.39 (1.14) |
| Self-harm | 3.49 (2.11–5.76) | 6.44 (3.65) | 2.41 (1.74–3.35) | 4.25 (2.86) |
| Non-elective hospitalisation | 1.17 (1.14–1.21) | 1.62 (1.54) | 1.15 (1.12–1.17) | 1.55 (1.48) |
| Nursing home admission | 3.11 (2.81–3.45) | 5.67 (5.06) | 2.14 (2.00–2.30) | 3.71 (3.41) |
| **Unconditional parametric survival regression** |  |  |  |  |
| All-cause mortality | 1.62 (1.51–1.74) | 2.62 (2.38) | 1.35 (1.29–1.41) | 2.03 (1.89) |
| Acute cardiovascular event | 1.32 (1.23–1.42) | 1.98 (1.77) | 1.25 (1.19–1.32) | 1.82 (1.67) |
| Hip fracture | 1.47 (1.31–1.66) | 2.31 (1.94) | 1.37 (1.26–1.49) | 2.08 (1.83) |
| Pneumonia | 1.19 (1.09–1.29) | 1.66 (1.40) | 1.12 (1.06–1.19) | 1.49 (1.3) |
| Self-harm | 2.96 (1.95–4.47) | 5.36 (3.32) | 2.30 (1.72–3.07) | 4.02 (2.83) |
| Non-elective hospitalisation | 1.17 (1.15–1.20) | 1.62 (1.56) | 1.15 (1.13–1.17) | 1.56 (1.5) |
| Nursing home admission | 3.07 (2.83–3.32) | 5.58 (5.11) | 2.17 (2.05–2.30) | 3.77 (3.53) |
| **Stratified Cox proportional hazard regression** |  |  |  |  |
| All-cause mortality | 1.66 (1.53–1.80) | 2.70 (2.43) | 1.36 (1.29–1.43) | 2.05 (1.89) |
| Acute cardiovascular event | 1.37 (1.27–1.48) | 2.09 (1.86) | 1.30 (1.23–1.37) | 1.92 (1.75) |
| Hip fracture | 1.51 (1.33–1.72) | 2.40 (1.99) | 1.37 (1.25–1.50) | 2.09 (1.82) |
| Pneumonia | 1.16 (1.06–1.28) | 1.59 (1.30) | 1.12 (1.04–1.20) | 1.48 (1.25) |
| Self-harm | 3.56 (2.14–5.91) | 6.57 (3.70) | 2.69 (1.90–3.81) | 4.82 (3.20) |
| Non-elective hospitalisation | 1.18 (1.15–1.22) | 1.65 (1.57) | 1.16 (1.13–1.19) | 1.59 (1.52) |
| Nursing home admission | 3.10 (2.80–3.44) | 5.66 (5.05) | 2.30 (2.14–2.48) | 4.03 (3.70) |

<sup>a</sup> Incidence rate ratios were adjusted on the same covariates as those reported in Tables 2 and 3. These estimates are identical to those reported in Supplementary Table S10.
<sup>b</sup> E-values for the point estimates are reported together with the e-values for the lower limit of the 95% confidence interval.
Interpretation: we found that the observed incidence rate ratio (IRR) for all-cause mortality of 1.66 could be fully explained away by an unmeasured confounder that was associated with both spousal loss and risk of death by an IRR of 2.70 each, above and beyond the measured confounders, but a weaker confounder could not do so. The lower limit of the confidence interval (namely, 1.53) could be moved to include the null by an unmeasured confounder that was associated with both spousal loss and death by a risk ratio of 2.43 each, above and beyond the measured confounders, but weaker confounding could not do so. Reference:
Linden A, Mathur MB, VanderWeele TJ. Conducting sensitivity analysis for unmeasured confounding in observational studies using E-values: The *eval* package. *Stata J* 2020;20:162–75. doi:10.1177/1536867X20909696. It should be noted that our analyses can also easily be replicated by using the online E-value calculator https://eval.evalue.hmdc.harvard.edu/app/

**Supplementary Table S21.** Sensitivity analysis: incidence rate ratios and 95% confidence intervals for acute stroke, acute myocardial infarction, pulmonary embolism, and aortic aneurysm or dissection after spousal loss

|  | **First year of follow-up** |  | **Complete follow-up** |  |
| --- | --- | --- | --- | --- |
|  | **Bereaved cases** | **Married controls** | **Bereaved cases** | **Married controls** |
| **Acute stroke** |  |  |  |  |
| No. of persons with the event | 744 | 561 | 1363 | 1172 |
| Person years of follow-up | 41 591 | 42 119 | 80 682 | 82 422 |
| Rate per 1000 person-years (95% CI) | 17.9 (16.6–19.2) | 13.3 (12.3–14.5) | 16.9 (16.0–17.8) | 14.2 (13.4–15.1) |
| Unadjusted incidence rate ratio (95% CI)<sup>a</sup> | 1.34 (1.20–1.50) |  | 1.17 (1.09–1.27) |  |
| Adjusted incidence rate ratio (95% CI)<sup>b</sup> | 1.36 (1.21–1.53) |  | 1.18 (1.08–1.29) |  |
| **Acute myocardial infarction** |  |  |  |  |
| No. of persons with the event | 599 | 496 | 1171 | 966 |
| Person years of follow-up | 41 643 | 42 148 | 80 927 | 82 572 |
| Rate per 1000 person-years (95% CI) | 14.4 (13.3–15.6) | 11.8 (10.8–12.9) | 14.5 (13.7–15.3) | 11.7 (11–12.5) |
| Unadjusted incidence rate ratio (95% CI)<sup>a</sup> | 1.22 (1.08–1.37) |  | 1.23 (1.12–1.34) |  |
| Adjusted incidence rate ratio (95% CI)<sup>b</sup> | 1.14 (1.00–1.30) |  | 1.18 (1.07–1.29) |  |
| **Pulmonary embolism** |  |  |  |  |
| No. of persons with the event | 227 | 171 | 419 | 317 |
| Person years of follow-up | 41 749 | 42 274 | 81 368 | 83 055 |
| Rate per 1000 person-years (95% CI) | 5.4 (4.8–6.2) | 4.0 (3.5–4.7) | 5.1 (4.7–5.7) | 3.8 (3.4–4.3) |
| Unadjusted incidence rate ratio (95% CI)<sup>a</sup> | 1.33 (1.09–1.62) |  | 1.33 (1.15–1.54) |  |
| Adjusted incidence rate ratio (95% CI)<sup>b</sup> | 1.31 (1.06–1.62) |  | 1.36 (1.17–1.60) |  |
| **Aortic aneurysm or dissection** |  |  |  |  |
| No. of persons with the event | 159 | 98 | 271 | 190 |
| Person years of follow-up | 41 776 | 42 294 | 81 459 | 83 152 |
| Rate per 1000 person-years (95% CI) | 3.8 (3.3–4.4) | 2.3 (1.9–2.8) | 3.3 (3.0–3.7) | 2.3 (2.0–2.6) |
| Unadjusted incidence rate ratio (95% CI)<sup>a</sup> | 1.62 (1.26–2.09) |  | 1.43 (1.19–1.72) |  |
| Adjusted incidence rate ratio (95% CI)<sup>b</sup> | 1.79 (1.27–2.52) |  | 1.47 (1.16–1.86) |  |

*Incidence rate ratios were calculated with conditional fixed-effect Poisson regression models grouped on the matched sets (sex and age), and further adjusted for diabetes, chronic heart failure, history of ischemic heart disease, history of cerebrovascular disease, atrial fibrillation, cardiac valve disease, peripheral vascular disease, other cardiovascular disease, number of other chronic comorbidities at baseline, and level of education. Further adjustment on cardiovascular medication use at baseline (antihypertensives, statins, and antithrombotic drugs) did not change the results.*

**Supplementary Table S22.** Sensitivity analysis: incidence rate ratios and 95% confidence intervals for acute cardiovascular events (broader definition) after spousal loss

|  | First year of follow-up |  | Complete follow-up |  |
| --- | --- | --- | --- | --- |
|  | Cases | Controls | Cases | Controls |
| **Bereaved cases vs. Married controls** |  |  |  |  |
| No. of persons with the event<sup>a</sup> | 2468 | 1884 | 4422 | 3628 |
| Person years of follow-up | 40 978 | 41 623 | 78 520 | 80 553 |
| Rate per 1000 person-years (95% CI) | 60.2 (57.9–62.7) | 45.3 (43.3–47.4) | 56.3 (54.7–58.0) | 45.0 (43.6–46.5) |
| Unadjusted incidence rate ratio (95% CI)<sup>b</sup> | 1.34 (1.26–1.43) |  | 1.26 (1.20–1.32) |  |
| Adjusted incidence rate ratio (95% CI)<sup>c</sup> | 1.34 (1.26–1.43) |  | 1.25 (1.19–1.31) |  |
| **Bereaved cases vs. Long-time widowed controls** |  |  |  |  |
| No. of persons with the event<sup>a</sup> | 2468 | 2141 | 4422 | 4106 |
| Person years of follow-up | 40 978 | 41 400 | 78 520 | 79 857 |
| Rate per 1000 person-years (95% CI) | 60.2 (57.9–62.7) | 51.7 (49.6–54.0) | 56.3 (54.7–58.0) | 51.4 (49.9–53.0) |
| Unadjusted incidence rate ratio (95% CI)<sup>b</sup> | 1.14 (1.08–1.21) |  | 1.08 (1.03–1.13) |  |
| Adjusted incidence rate ratio (95% CI)<sup>c</sup> | 1.16 (1.09–1.24) |  | 1.09 (1.04–1.15) |  |

<sup>a</sup> The “broader” definition of acute cardiovascular events is detailed in Supplementary Table S3.
<sup>b</sup> Unadjusted conditional fixed-effect Poisson regression model with grouping on the matched sets (sex and age).
<sup>c</sup> Conditional fixed-effect Poisson regression model with grouping on the matched sets (sex and age), adjusted for diabetes, chronic heart failure, history of ischemic heart disease, history of cerebrovascular disease, atrial fibrillation, cardiac valve disease, peripheral vascular disease, other cardiovascular disease, number of other chronic comorbidities at baseline, and level of education.

**Supplementary Table S23.** Sensitivity analysis: incidence rate ratios and 95% confidence intervals for injurious falls after spousal loss

|  | First year of follow-up |  | Complete follow-up |  |
| --- | --- | --- | --- | --- |
|  | Cases | Controls | Cases | Controls |
| **Bereaved cases vs. Married controls** |  |  |  |  |
| No. of persons with the event<sup>a</sup> | 2968 | 2338 | 5387 | 4469 |
| Person years of follow-up | 40 414 | 41 226 | 76 265 | 78 878 |
| Rate per 1000 person-years (95% CI) | 73.4 (70.8–76.1) | 56.7 (54.5–59.1) | 70.6 (68.8–72.5) | 56.7 (55.0–58.3) |
| Unadjusted incidence rate ratio (95% CI)<sup>b</sup> | 1.28 (1.21–1.36) |  | 1.24 (1.19–1.29) |  |
| Adjusted incidence rate ratio (95% CI)<sup>c</sup> | 1.28 (1.21–1.36) |  | 1.24 (1.18–1.29) |  |
| **Bereaved cases vs. Long-time widowed controls** |  |  |  |  |
| No. of persons with the event<sup>a</sup> | 2968 | 2653 | 5387 | 5058 |
| Person years of follow-up | 40 414 | 40 984 | 76 265 | 77955 |
| Rate per 1000 person-years (95% CI) | 73.4 (70.8–76.1) | 64.7 (62.3–67.2) | 70.6 (68.8–72.5) | 64.9 (63.1–66.7) |
| Unadjusted incidence rate ratio (95% CI)<sup>b</sup> | 1.12 (1.07–1.19) |  | 1.07 (1.03–1.12) |  |
| Adjusted incidence rate ratio (95% CI)<sup>c</sup> | 1.13 (1.07–1.20) |  | 1.08 (1.04–1.13) |  |

<sup>a</sup> Unplanned hospitalisation for fall (ICD-10 codes W00-W19).
<sup>b</sup> Unadjusted conditional fixed-effect Poisson regression model with grouping on the matched sets (sex and age).
<sup>c</sup> Conditional fixed-effect Poisson regression model with grouping on the matched sets (sex and age), adjusted for osteoporosis, osteoarthritis, Parkinson's disease, dementia, number of other chronic comorbidities at baseline, history of fall-related injury and history of alcohol-related disease in the past 5 years.

**Supplementary Table S24.** Post-hoc analysis: incidence rate ratios and 95% confidence intervals for all-cause mortality and adverse health events during the year after spousal loss, according to the illness trajectory of the deceased spouse

|  | Illness trajectory of the deceased spouse |  |  |  |
| --- | --- | --- | --- | --- |
|  | Cancer | Organ failure | Prolonged dwindling | Sudden death |
| **No. of bereaved cases** | 17 082 | 14 359 | 8265 | 2968 |
| **All-cause mortality** |  |  |  |  |
| Incidence rate among bereaved cases, per 1000 (95% CI) | 38.2 (35.3–41.3) | 56.1 (52.4–60.2) | 56.9 (51.9–62.3) | 57.5 (49.4–67.0) |
| Incidence rate among married controls, per 1000 (95% CI) | 26.5 (24.2–29.1) | 32.4 (29.5–35.5) | 38.6 (34.6–43.1) | 25.3 (20.2–31.8) |
| Unadjusted IRR (95% CI)<sup>a</sup> | 1.47 (1.30–1.67) | 1.76 (1.56–1.98) | 1.49 (1.28–1.72) | 2.46 (1.84–3.27) |
| Adjusted IRR (95% CI)<sup>b</sup> | 1.52 (1.32–1.76) | 1.82 (1.59–2.09) | 1.45 (1.23–1.71) | 2.82 (1.99–4.01) |
| **Acute cardiovascular event** |  |  |  |  |
| Incidence rate among bereaved cases, per 1000 (95% CI) | 37.9 (35.1–41.0) | 47.7 (44.2–51.5) | 52.8 (48.0–58.1) | 41.0 (34.1–49.1) |
| Incidence rate among married controls, per 1000 (95% CI) | 30.2 (27.7–32.9) | 33.2 (30.4–36.4) | 38.5 (34.4–43.1) | 36.5 (30.2–44.2) |
| Unadjusted incidence rate ratio (95% CI)<sup>a</sup> | 1.26 (1.12–1.42) | 1.44 (1.28–1.63) | 1.38 (1.19–1.61) | 1.15 (0.88–1.50) |
| Adjusted incidence rate ratio (95% CI)<sup>b</sup> | 1.26 (1.11–1.44) | 1.42 (1.25–1.62) | 1.42 (1.20–1.66) | 1.11 (0.83–1.49) |
| **Hip fracture** |  |  |  |  |
| Incidence rate among bereaved cases, per 1000 (95% CI) | 11.6 (10.1–13.4) | 18.2 (16.1–20.6) | 18.9 (16.2–22.2) | 15.8 (11.8–21.2) |
| Incidence rate among married controls, per 1000 (95% CI) | 9.5 (8.2–11.1) | 11.4 (9.8–13.4) | 11.9 (9.7–14.5) | 10.3 (7.2–14.8) |
| Unadjusted incidence rate ratio (95% CI)<sup>a</sup> | 1.21 (0.98–1.49) | 1.56 (1.28–1.91) | 1.58 (1.22–2.04) | 1.50 (0.95–2.38) |
| Adjusted incidence rate ratio (95% CI)<sup>b</sup> | 1.20 (0.95–1.50) | 1.63 (1.32–2.00) | 1.61 (1.22–2.13) | 1.89 (1.09–3.29) |
| **Pneumonia** |  |  |  |  |
| Incidence rate among bereaved cases, per 1000 (95% CI) | 24.5 (22.2–27.0) | 30.6 (27.8–33.6) | 31.6 (27.9–35.8) | 24.3 (19.2–30.8) |
| Incidence rate among married controls, per 1000 (95% CI) | 20.1 (18.1–22.4) | 25.8 (23.3–28.6) | 28.0 (24.5–31.9) | 24.2 (19.1–30.6) |
| Unadjusted incidence rate ratio (95% CI)<sup>a</sup> | 1.21 (1.04–1.40) | 1.18 (1.03–1.36) | 1.14 (0.95–1.37) | 1.02 (0.72–1.42) |
| Adjusted incidence rate ratio (95% CI)<sup>b</sup> | 1.20 (1.02–1.42) | 1.12 (0.97–1.30) | 1.14 (0.93–1.39) | 1.03 (0.69–1.52) |
| **Self-harm** |  |  |  |  |
| Incidence rate among bereaved cases, per 1000 (95% CI) | 2.0 (1.5–2.8) | 1.8 (1.2–2.7) | 2.2 (1.4–3.6) | 4.2 (2.4–7.4) |
| Incidence rate among married controls, per 1000 (95% CI) | 0.8 (0.4–1.3) | 0.5 (0.2–1.0) | 0.7 (0.3–1.6) | 1.4 (0.5–3.7) |
| Unadjusted incidence rate ratio (95% CI)<sup>a</sup> | 2.62 (1.38–4.96) | 3.57 (1.55–8.26) | 3.00 (1.19–7.56) | 3.00 (0.97–9.30) |
| Adjusted incidence rate ratio (95% CI)<sup>b</sup> | 3.52 (1.60–7.76) | 4.84 (1.58–14.84) | 3.97 (1.14–13.84) | 1.95 (0.57–6.62) |

<sup>a</sup> Unadjusted conditional fixed-effect Poisson regression model with grouping on the matched sets (sex and age). Reference group: married controls.
<sup>b</sup> Conditional fixed-effect Poisson regression model with grouping on the matched sets (sex and age) and further adjusted on the same covariates as those reported in Table 2

**Supplementary Table S25.** Post-hoc analysis: incidence rate ratios and 95% confidence intervals for all-cause mortality after spousal loss, according to Hospital Frailty Risk Score at baseline

|  | First year of follow-up |  | Complete follow-up |  |
| --- | --- | --- | --- | --- |
|  | Bereaved cases | Married controls | Bereaved cases | Married controls |
| **Low risk of frailty (<5)** |  |  |  |  |
| No. of deaths | 1080 | 697 | 2387 | 1885 |
| Person years of follow-up | 33 724 | 34 173 | 66 396 | 67 709 |
| Rate per 1000 person-years (95% CI) | 32.0 (30.2–34.0) | 20.4 (18.9–22.0) | 36.0 (34.5–37.4) | 27.8 (26.6–29.1) |
| Unadjusted incidence rate ratio (95% CI)<sup>a</sup> | 1.57 (1.43–1.73) |  | 1.29 (1.22–1.37) |  |
| Adjusted incidence rate ratio (95% CI)<sup>b</sup> | 1.55 (1.41–1.71) |  | 1.28 (1.20–1.36) |  |
| **Intermediate risk of frailty (5-9)** |  |  |  |  |
| No. of deaths | 557 | 306 | 1037 | 714 |
| Person years of follow-up | 5789 | 5735 | 11 023 | 11 104 |
| Rate per 1000 person-years (95% CI) | 96.2 (88.6–104.6) | 53.4 (47.7–59.7) | 94.1 (88.5–100.0) | 64.3 (59.8–69.2) |
| Unadjusted incidence rate ratio (95% CI)<sup>a</sup> | 1.80 (1.57–2.07) |  | 1.46 (1.33–1.61) |  |
| Adjusted incidence rate ratio (95% CI)<sup>b</sup> | 1.86 (1.61–2.14) |  | 1.52 (1.38–1.67) |  |
| **High risk of frailty (≥10)** |  |  |  |  |
| No. of deaths | 423 | 303 | 767 | 630 |
| Person years of follow-up | 2353 | 2452 | 4253 | 4471 |
| Rate per 1000 person-years (95% CI) | 179.8 (163.4–197.8) | 123.6 (110.4–138.3) | 180.4 (168.0–193.6) | 140.9 (130.3–152.3) |
| Unadjusted incidence rate ratio (95% CI)<sup>a</sup> | 1.45 (1.26–1.69) |  | 1.28 (1.15–1.42) |  |
| Adjusted incidence rate ratio (95% CI)<sup>b</sup> | 1.51 (1.30–1.76) |  | 1.34 (1.20–1.49) |  |

<sup>a</sup> Unadjusted incidence rate ratio were estimated by unconditional Poisson regression analysis
<sup>b</sup> Adjusted incidence rate ratio were estimated by unconditional Poisson regression analysis with adjustment on sex, age, level of education, chronic obstructive pulmonary disease, diabetes, chronic heart failure, history of ischemic heart diseases and history of solid cancer in the previous 5 years, and number of other chronic diseases

**Supplementary Figure S7.**
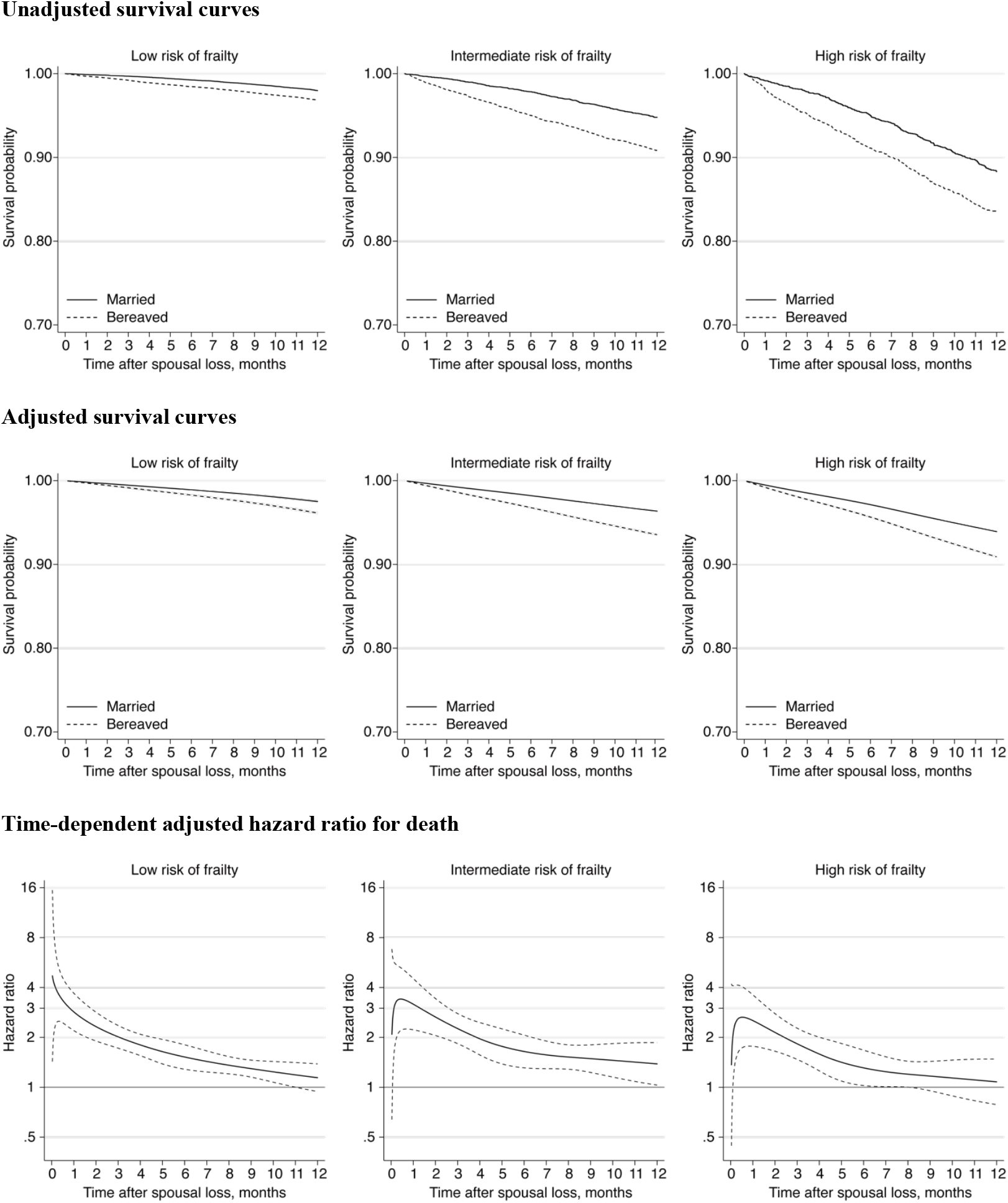
Post-hoc analysis: survival of bereaved cases and married controls and hazard ratio for death during the first year after spousal loss, according to frailty risk at baseline. Unadjusted survival curves were produced by using Kaplan-Meier (non-parametric) estimators. Adjusted survival curves were produced by fitting Royston-Parmar flexible parametric survival regression models adjusted for sex, age, chronic obstructive pulmonary disease, diabetes, chronic heart failure, history of ischemic heart diseases, history of solid cancer in the previous 5 years, and number of other chronic diseases. Curves representing the hazard ratio for death as a function of time since spousal loss were obtained by fitting flexible parametric survival regression models with time-dependent effects using restricted cubic splines.

**Supplementary Table S26.** Post-hoc analysis: competing-risks regression models

|  | **Flexible parametric survival**<br>**model without competing risk<sup>a</sup>** | **Fine-Gray competing risk**<br>**models<sup>b</sup>** | **Flexible parametric competing-**<br>**risks regression models<sup>c</sup>** |
| --- | --- | --- | --- |
|  | Unadjusted HR (95% CI) | Unadjusted SHR (95% CI) | Unadjusted SHR (95% CI) |
| **First year of follow-up** |  |  |  |
| Acute cardiovascular event | 1.33 (1.24–1.42) | 1.32 (1.23–1.41) | 1.32 (1.23–1.41) |
| Hip fracture | 1.45 (1.29–1.64) | 1.44 (1.28–1.62) | 1.44 (1.28–1.62) |
| Pneumonia | 1.17 (1.07–1.27) | 1.16 (1.06–1.26) | 1.16 (1.06–1.26) |
| Self-harm | 3.00 (1.98–4.53) | 2.97 (1.96–4.49) | 2.97 (1.96–4.49) |
| ER visit or non-elective hospitalisation | 1.16 (1.14–1.19) | 1.15 (1.13–1.18) | 1.15 (1.13–1.18) |
| Nursing home admission | 2.63 (2.43–2.84) | 2.61 (2.41–2.82) | 2.60 (2.41–2.81) |
| **Complete follow-up** |  |  |  |
| Acute cardiovascular event | 1.25 (1.19–1.32) | 1.24 (1.18–1.30) | 1.24 (1.18–1.30) |
| Hip fracture | 1.35 (1.24–1.46) | 1.32 (1.22–1.44) | 1.32 (1.21–1.44) |
| Pneumonia | 1.10 (1.03–1.17) | 1.08 (1.02–1.15) | 1.08 (1.02–1.15) |
| Self-harm | 2.31 (1.73–3.09) | 2.28 (1.71–3.05) | 2.28 (1.70–3.04) |
| ER visit or non-elective hospitalisation | 1.14 (1.11–1.16) | 1.13 (1.11–1.15) | 1.13 (1.10–1.15) |
| Nursing home admission | 1.87 (1.77–1.98) | 1.85 (1.75–1.95) | 1.85 (1.75–1.95) |

Abbreviations: HR, hazard ratio; SHR, subdistribution hazard ratio; CI, confidence interval; ER, emergency room.
<sup>a</sup>Flexible parametric survival regression models (Royston-Parmar). Individuals are censored at time of death. These were fit by using the `stpm2` command in Stata.
<sup>b</sup>Fine-Gray competing-risk regressions, which model the subhazard function of an event of interest in the presence of the competing risk of death from any other cause. Hazard ratios obtained from these models correspond to the relative change in the instantaneous rate at which the event of interest occurs among subjects who are currently event-free or who have experienced the competing event. When the event of interest is rare, the subdistribution hazard ratio can be interpreted as an approximation of the relative change in the cumulative incidence of that event. Fine-Gray competing-risks regressions were fit by using the `stcrreg` command in Stata.
<sup>c</sup>Flexible parametric competing-risks regression models, which were fit by using the `stpm2cr` command in Stata (Mozumder et al., 2017).

